# Frontiers of cervical cancer research: an analysis from the top 100 most influential articles in the field

**DOI:** 10.1101/2024.11.08.24316970

**Authors:** Lei Liang, Chuangxiu Song, Chun Chang, Shichao Chen, Bo Yang, Li Sun

**Affiliations:** The 980 Hospital of the Joint Logistic Support Force of the Chinese People’s Liberation Army. Shijiazhuang City, Hebei Province 050082, China; Neck-Shoulder and Lumbocrural Pain Hospital of Shandong First Medical University. Jinan City, Shandong Province 250355, China

**Keywords:** cervical cancer, cervical cancer screening, bibliometric analysis, CiteSpace, visualization, VOSviewer

## Abstract

**Backgroung:** Cervical cancer (CC) is one of the common gynecological malignancies, which has a serious negative impact on the quality of life of patients, and brings huge economic pressure and burden to society. The most effective treatment for CC has become an important issue worldwide.

**Method:** In order to determine the current research hotspots and development trends in a particular field, we conducted a bibliometric analysis of the top 100 cited papers. To achieve this, we searched for the top 100 most cited papers on the Science Citation Index Expanded (SCI Expanded) on the Web of Science (WOS), based on the CCS (citation count score). We then reviewed relevant literature from different years, countries/regions, institutions, authors, keywords, and references. Using VOS viewer and Cite Space software, we constructed a knowledge map, and finally compiled the relevant information we retrieved from the literature using Excel. Through this process, we were able to make predictions about the current focus and trends in the field.

**Results:** Between 2013 and 2023, numerous journals published the top 100 cited research papers. The International Agency for Research on Cancer and the American Cancer Society were the most cited institutions. The United States contributed the most papers, followed by France and England. The top 5 keyword co-occurrences were cervical cancer, women, human papillomavirus, United States, and colorectal cancer. Cluster analysis results suggest that future research in CCS may focus on prophylaxis and HPV vaccination.

**Conclusion:** Bibliometric analysis can quickly and intuitively identify the focus and boundaries of cervical cancer research. Our findings suggest that prophylaxis and HPV vaccination may be the focus and trend for future research on cervical cancer.

## 1. Introduction

Cervical cancer is one of the common female malignancies, and persistent infection with high-risk human papillomavirus (HPV) is the main cause of its development. Globally, cervical cancer is characterized by unequal incidence and mortality rates [1]. Data from the 2018 Global Cancer Registry report show that around 85% of cervical cancers occur in low- and middle-income countries, and that cervical cancer incidence in low- and middle-income countries is nearly twice the incidence of high-income countries, with a mortality rate of around three times that of high-income countries [2]. Without aggressive and effective measures, more than 44 million women in low- and middle-income countries will suffer from cervical cancer in the next 50 years. It significantly increases the economic burden on individuals and society, as well as the mental stress on patients and their families. Therefore, the prevention, control, and treatment of cervical cancer are of great significance to patients, families, and society [3]. Cervical cancer is currently the only cancer with a clear etiology that is preventable and curable, and with the three-tier prevention and control means of HPV vaccine, screening, and early diagnosis and treatment, it is expected to be the world’s first malignant tumor to be eliminated [4].

The first article on CCS was published in 2013 year [5]. In the past 10 years, researchers have made significant progress in the diagnosis and screening of CC. However, understanding the overall progress and research trends in the CC disease area is challenging, and scientific analyses using bibliometric techniques are essential [6–7]. Screening by understanding the top 100 cited articles in the field of CC will provide researchers with a deeper understanding of current research priorities. Bibliometric analyses can fully assist researchers in studying a particular current research area and is a very effective method of bibliometric analysis. Unfortunately, no studies with a similar focus were found in our search of the prior literature. The bibliometric analysis was used to identify the top 100 cited articles in the field and to analyze the research hotspots and research trends in the field. We believe that this study will help accelerate the development of the field of CCS research and encourage scholars to produce new findings in this area.

## 2. Materials and Methods

### 2.1. Search strategy

We used WoSCC literature related to managing cervical cancer screening against cervical cancer. The search strategy included cervical cancer screening and its synonyms (subject terms), and cervical cancer and its synonyms. The publication timeframe was from the build to December 2023 (search date: 01 January 2024). We finally included the top 100 most cited articles for the study. The specific search strategy is shown in

### 2.2. Inclusion criteria

Publications are ranked in descending order of overall citation frequency. Selected articles and reviews from different academic journals including Cervical Cancer Screening, and Cervical Malignancies. Letters, conference abstracts, book reviews, conference presentations, and news reports were excluded. Initial screening was done independently by two scholars to eliminate irrelevant topics. They identified the most cited publications by reading the titles, abstracts, keywords, and full text of 100 papers. If two researchers disagree on whether an article should be included, we must consult a third researcher to find a solution.

### 2.3. Bibliometric analysis

According to the system functionality of WoSCC, after downloading the details of country/region distribution, year of publication, institution, journal, authors, and keyword-related documents, we used Office 16 to sort and analyze the publications. CiteSpace 6.1.R6, VOSviewer 1.6.19, Scimago Graphica 1.0.30 Visual analyses of article features to identify current research hotspots and potential trends in the field of CCS. VOSviewer is a bibliometrics application for the creation of scientometric networks and knowledge visualization [8]. CiteSpace was used for identifying the network of co-authors of countries/institutions, bitmap analysis, and keyword detection with citation explosion [9]. Citation explosion indicates an increase in hot topics for potential work in the field over a given time, which is an important indicator for identifying research trends.

### 2.4. Bibliometric analysis terminology

Knowledge graph visualization is an outgrowth of the development of graphical data technologies. Data visualization is the aggregation of available information into a large network that contains semantic models of data for users to query and explore. In this way, raw data is converted into graphical information and the information is presented more intuitively and visually. Intermediate centrality is the number of times a node is located on the shortest path between other nodes and is one of the key metrics for measuring the importance of a node. Nodes with intermediate centrality greater than 0.1 are generally considered to be critical nodes. The more times a node lies on the shortest path, the higher the centrality of the node.

### 2.5 Ethical review

As this article is a visual analysis summary of previously published articles, it does not need to pass ethical review.

## 3. Results

**Citation characteristics of the included**

### 3.1 Articles

Between 1929 and 2023, 28,443 articles were published on early screening for CC. Two researchers combed through the research topics and article abstracts to exclude those articles in which CCS was not the main research topic. The eligible articles were added to this study in descending order according to the frequency of citation.

The top 100 cited papers are shown in Table 2. These 100 papers received a total of 56802 citations. There are 6,576 references in the top 100 cited papers. The most frequently cited article is “Global Cancer Statistics.2012” [10] by Torre, Lindsey A. (14567 citations). Next is “Cancer Statistics,2019” [11] by Siegel, RL (4436 citations), “Global Cancer Incidence and Mortality Rates and Trends-An Update” [12] (2501 citations), “The Global Burden of Cancer 2013 Global Burden of Disease Cancer Collaboration” [13] (2107 citations), “Estimates of incidence and mortality of cervical cancer in 2018: a worldwide analysis” [14] (1785 citations), “Global surveillance of cancer survival 1995-2009: analysis of individual data for 25,676,887 patients from 279 population-based registries in 67 countries (CONCORD-2)” [15] (1673 citations), “Cervical cancer” [1] (1202 citations), “Worldwide burden of cancer attributable to HPV by site, country and HPV type” [16] (1181 citations), “Efficacy of HPV-based screening for prevention of invasive cervical cancer: follow-up of four European randomised controlled trials” [17] (1139 citations), “Cancer statistics for African Americans, 2019” [18] (1116 citations). These are the articles that have been cited more than 1,000 times.

**Table 1.**
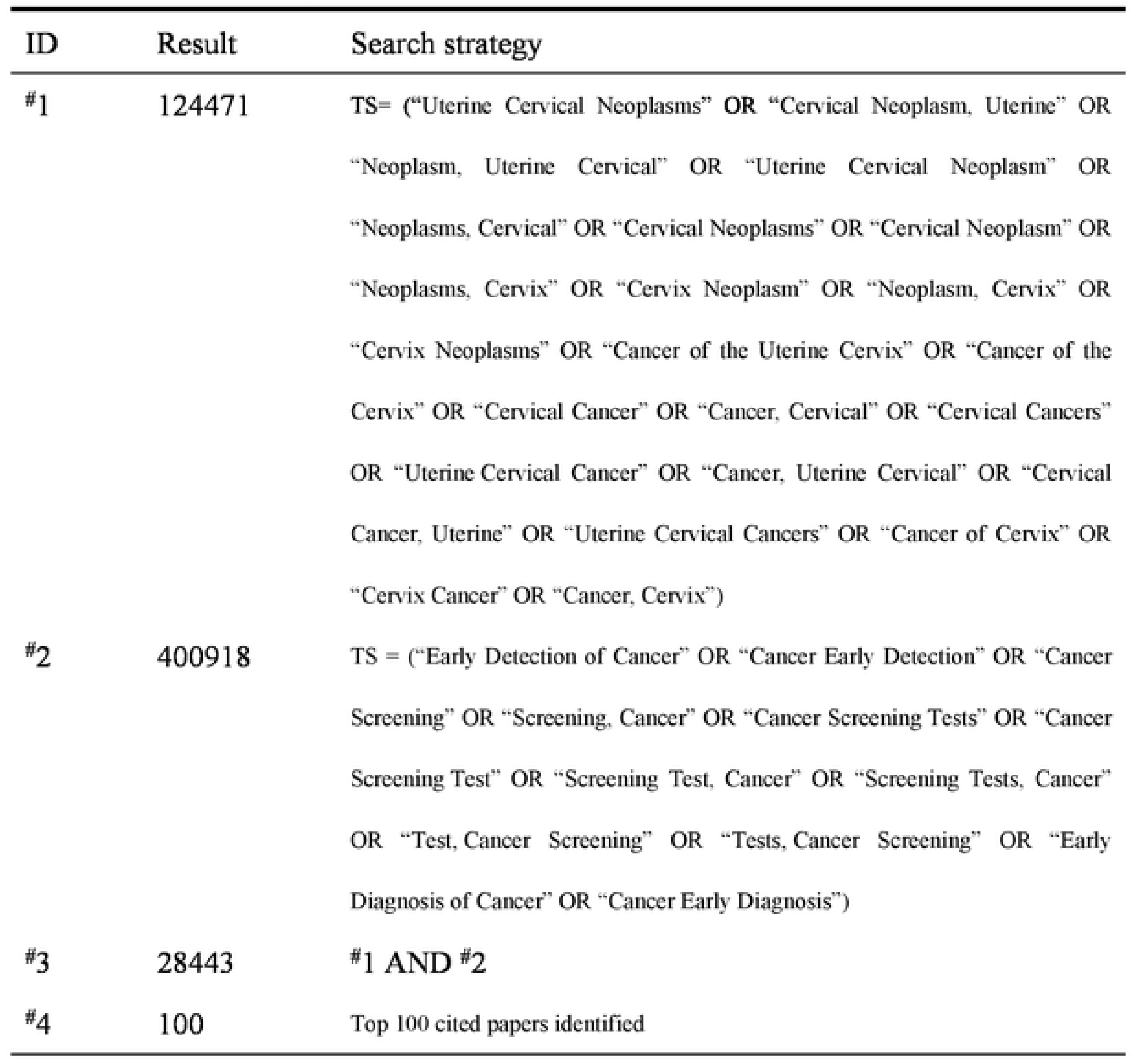
Search strategy.

**Table 2.**
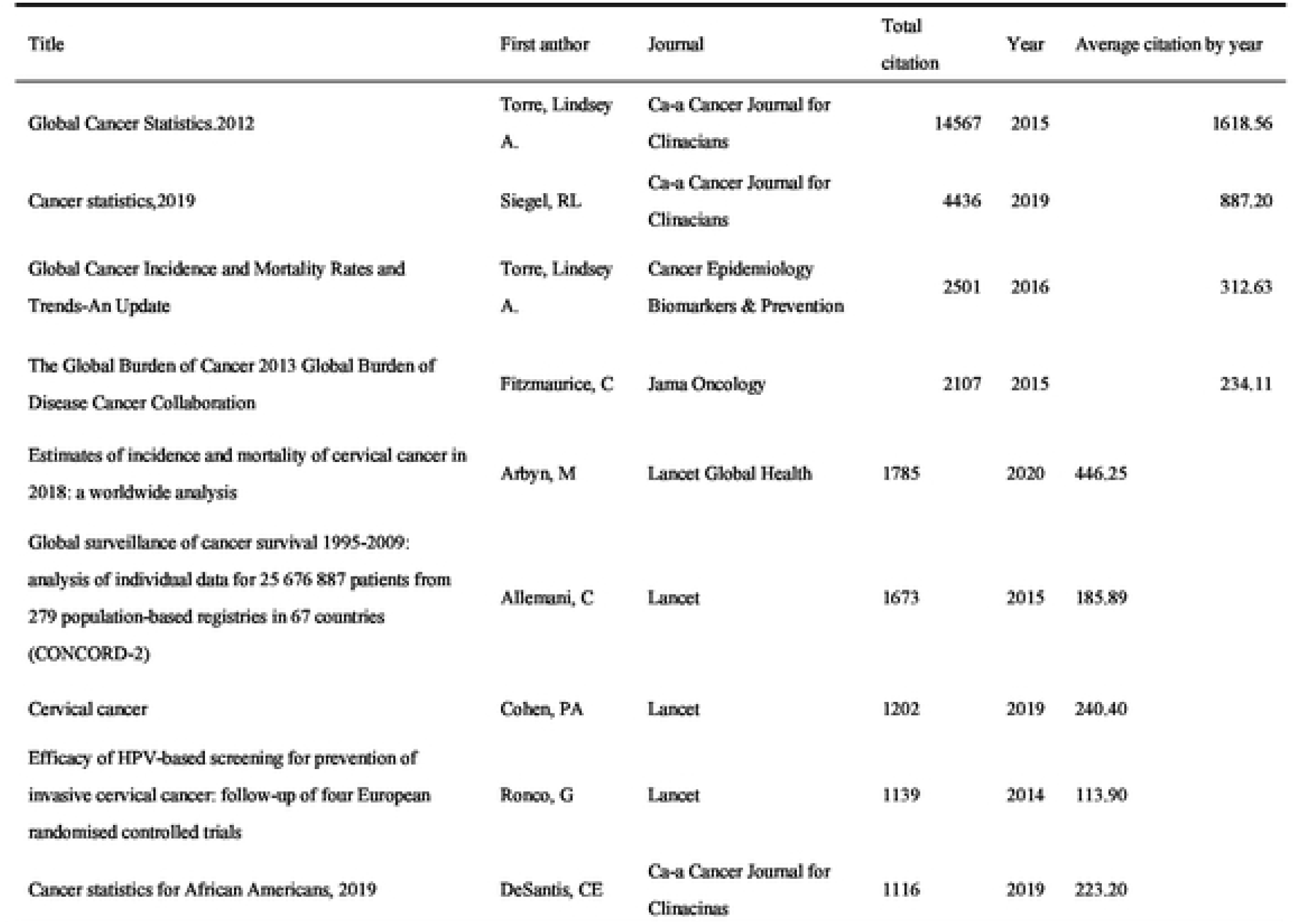
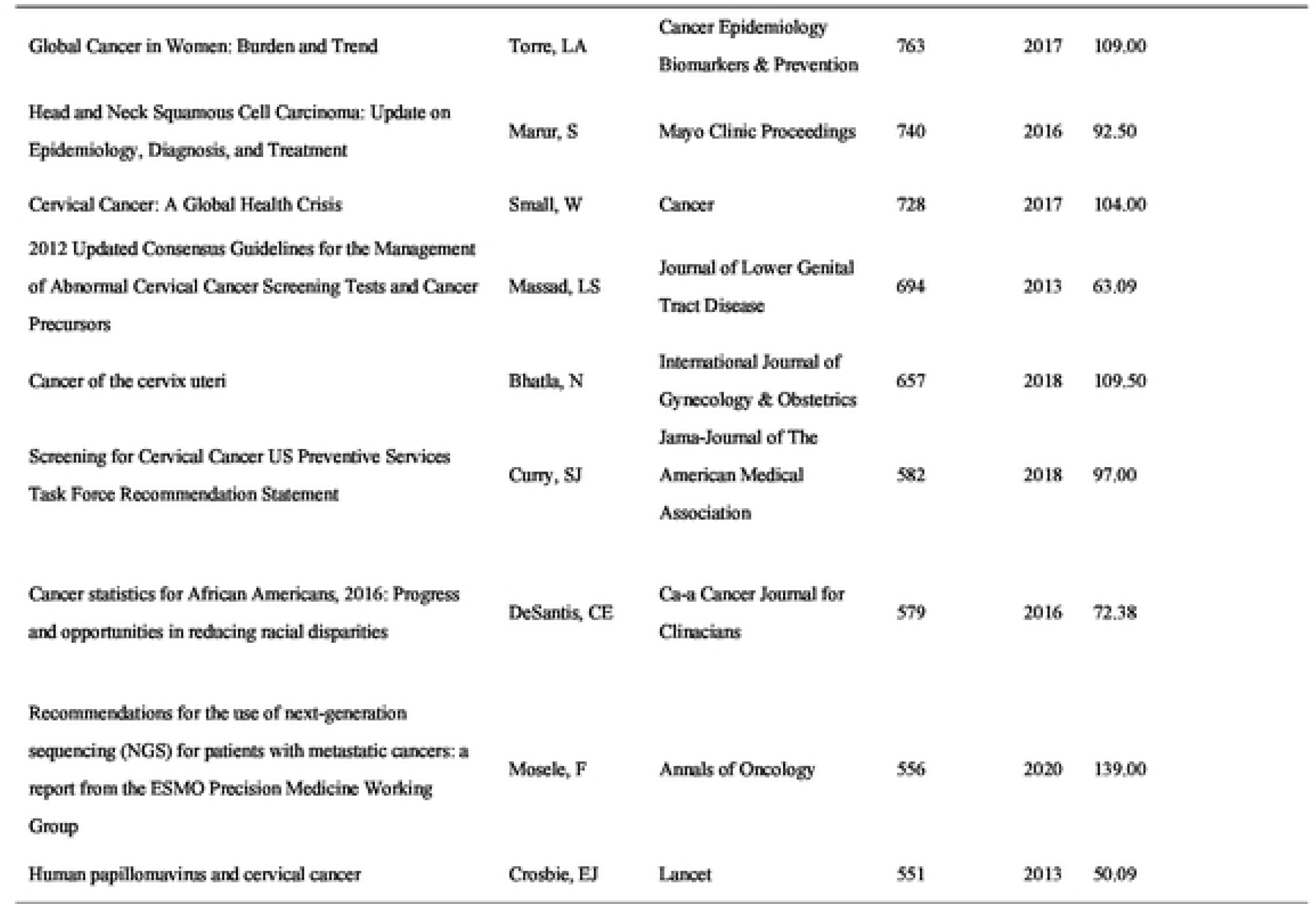
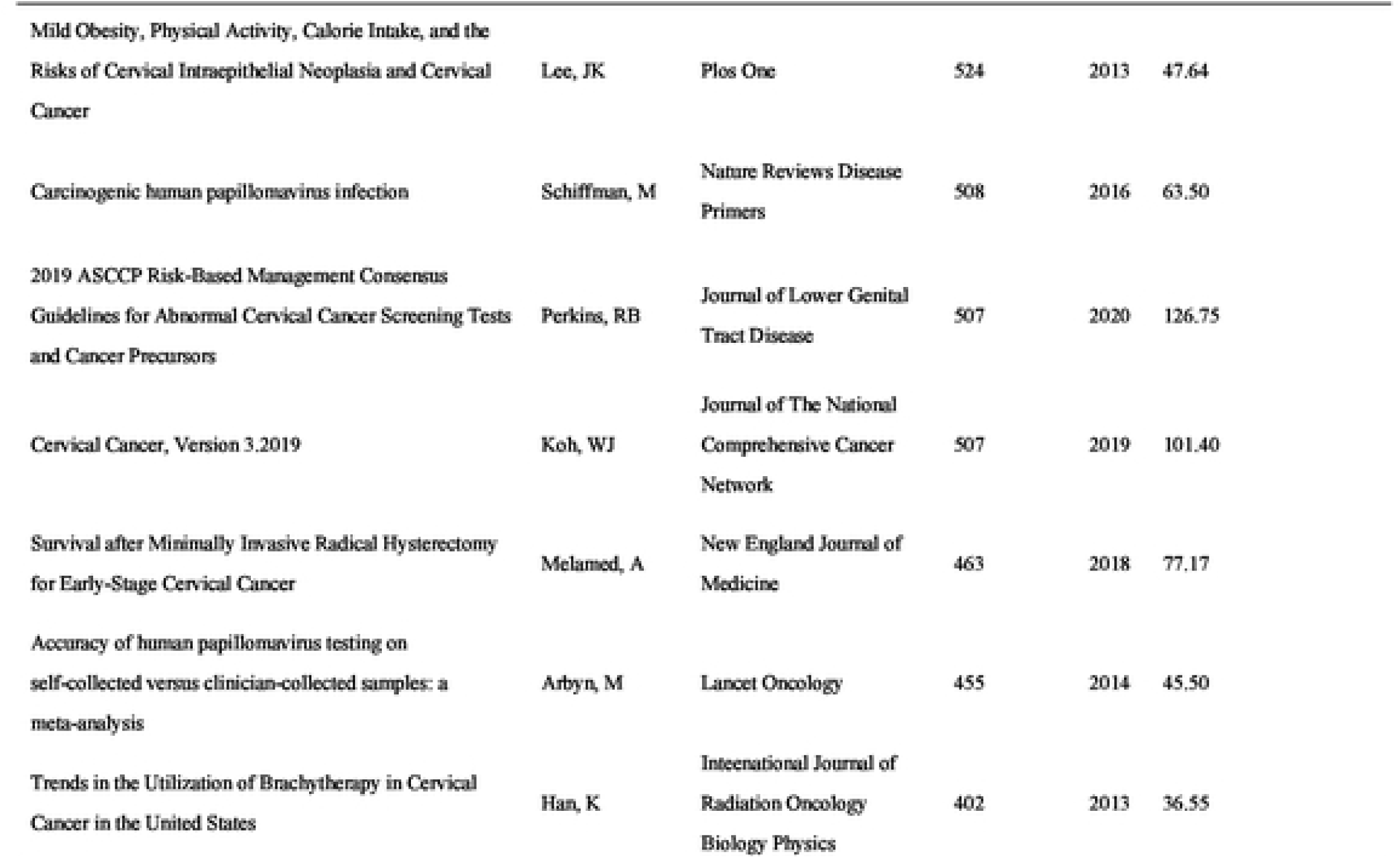
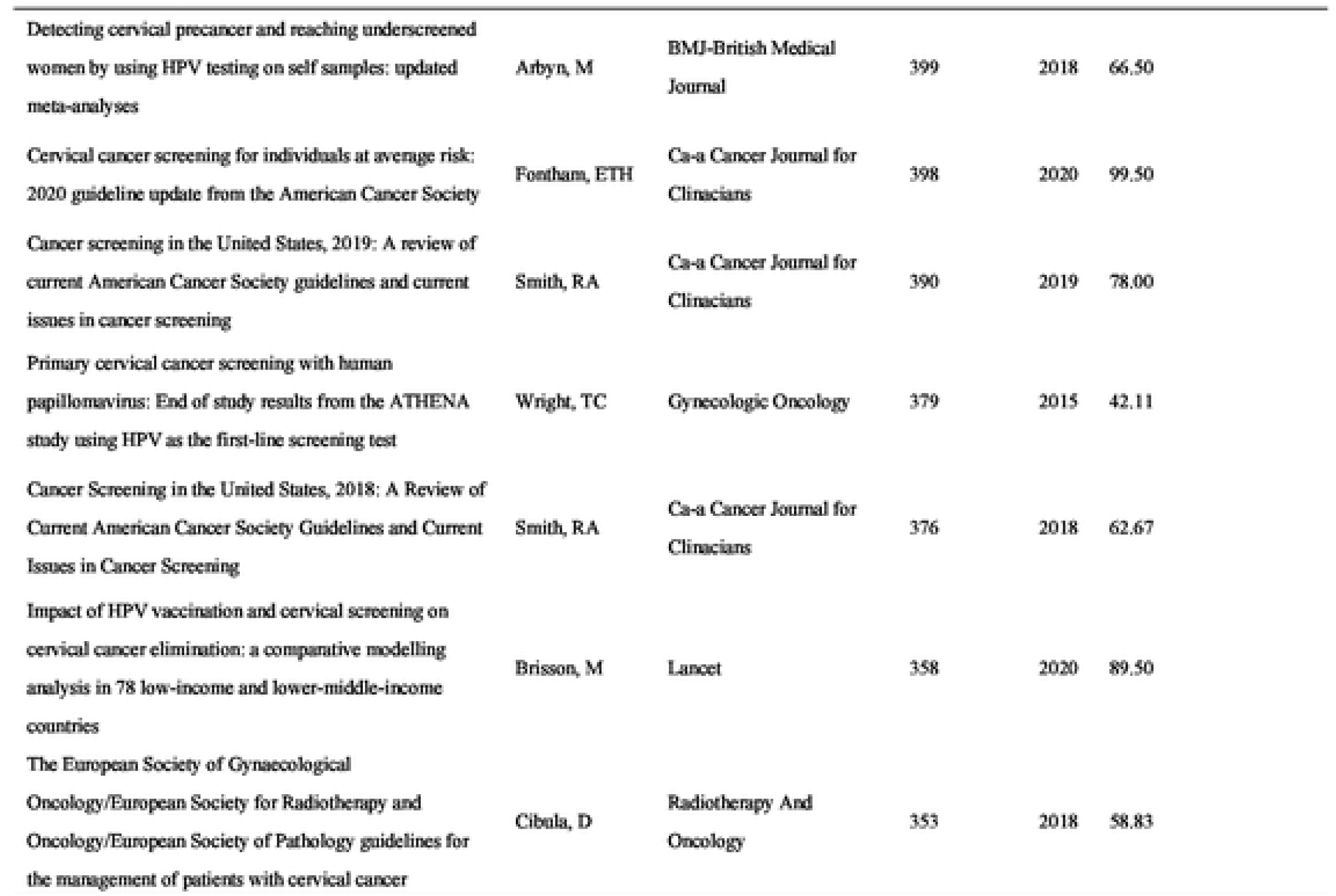
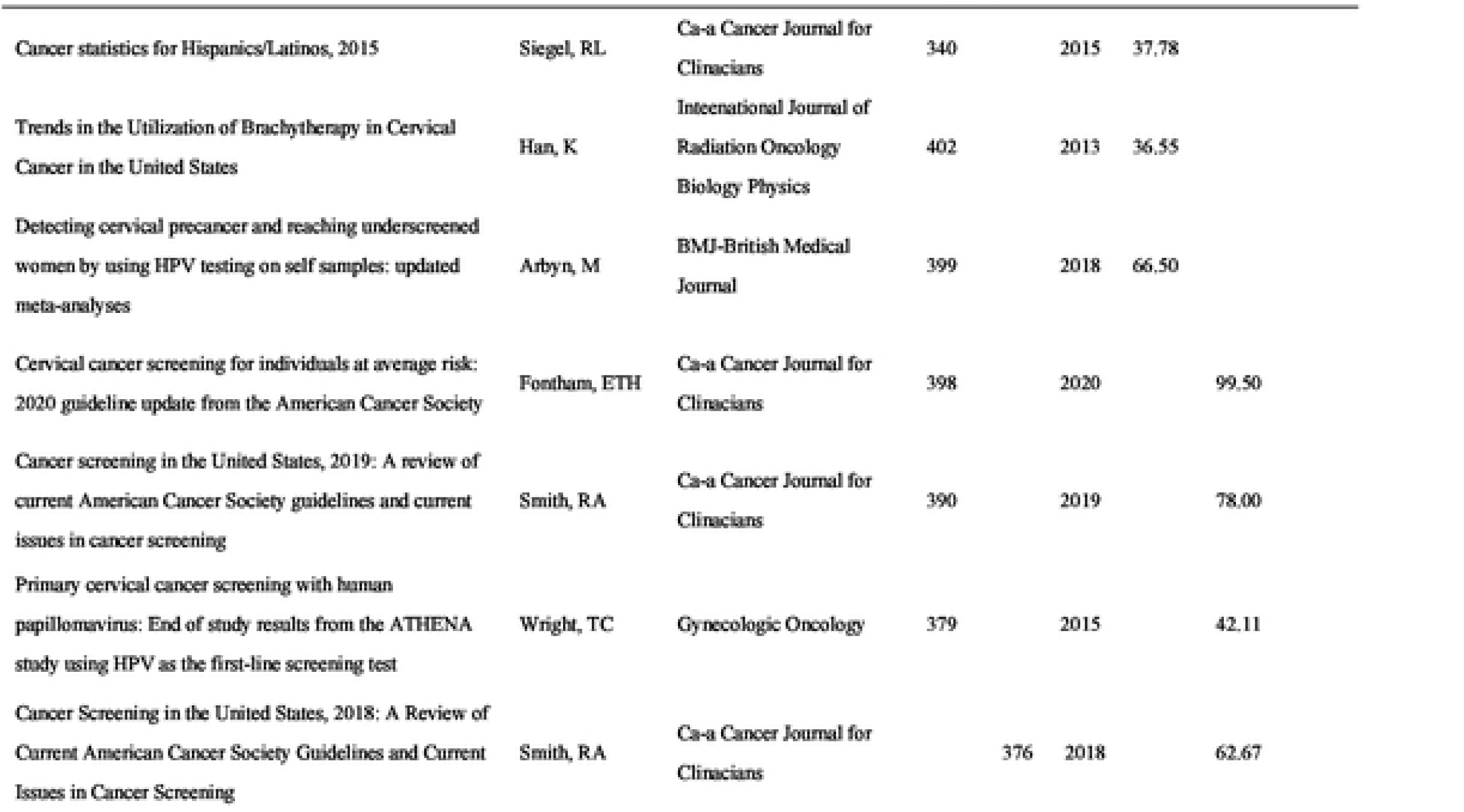
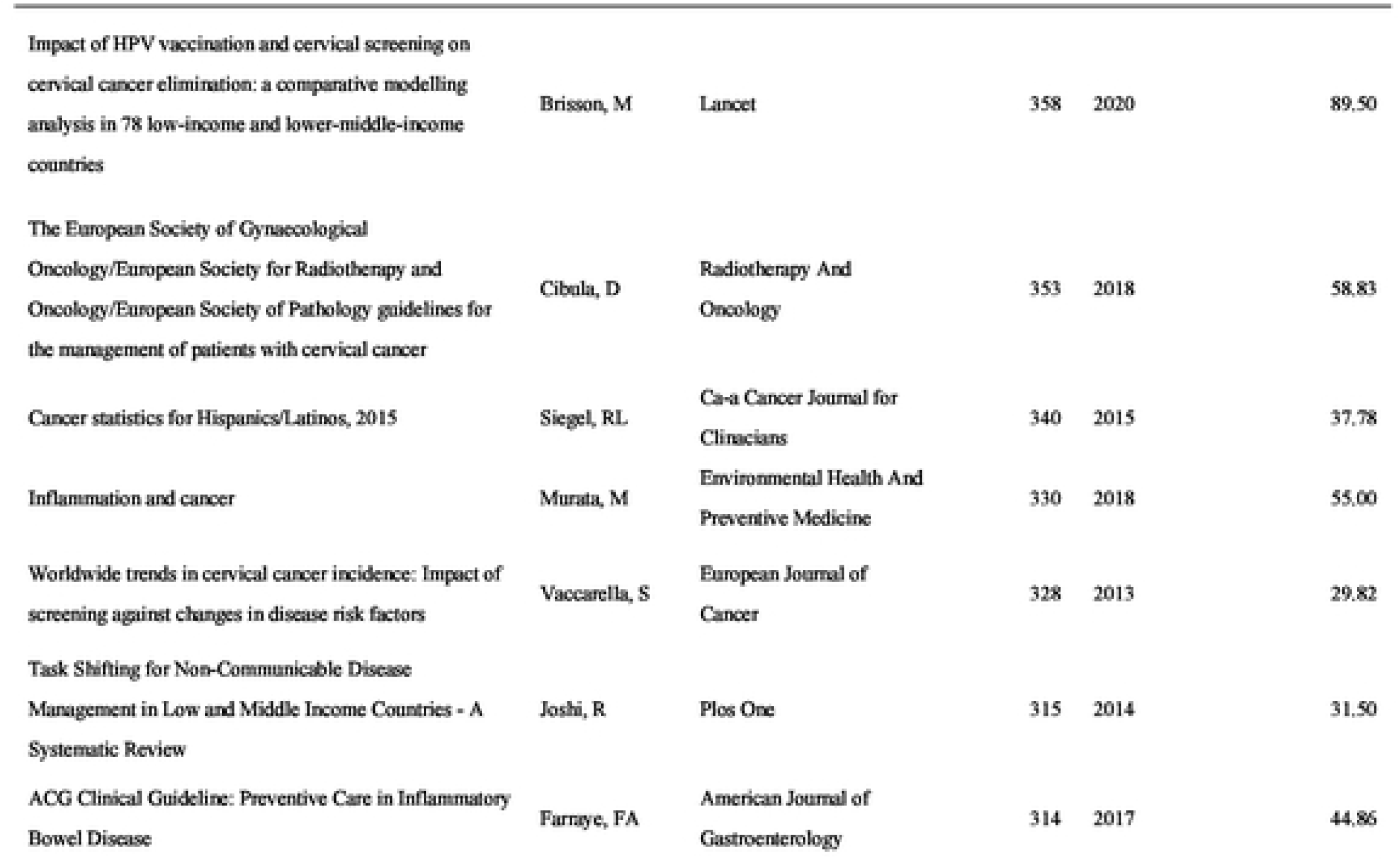
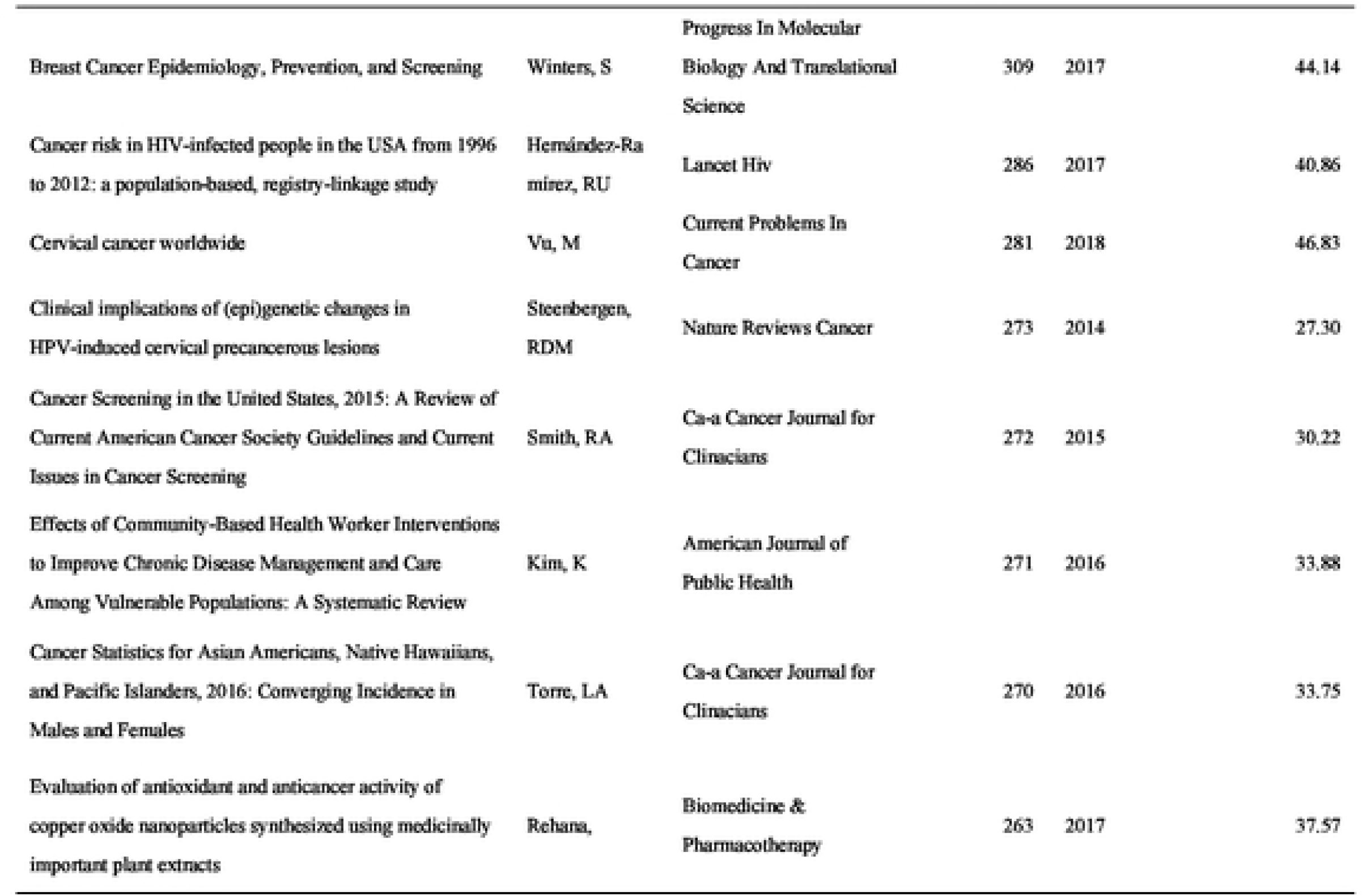
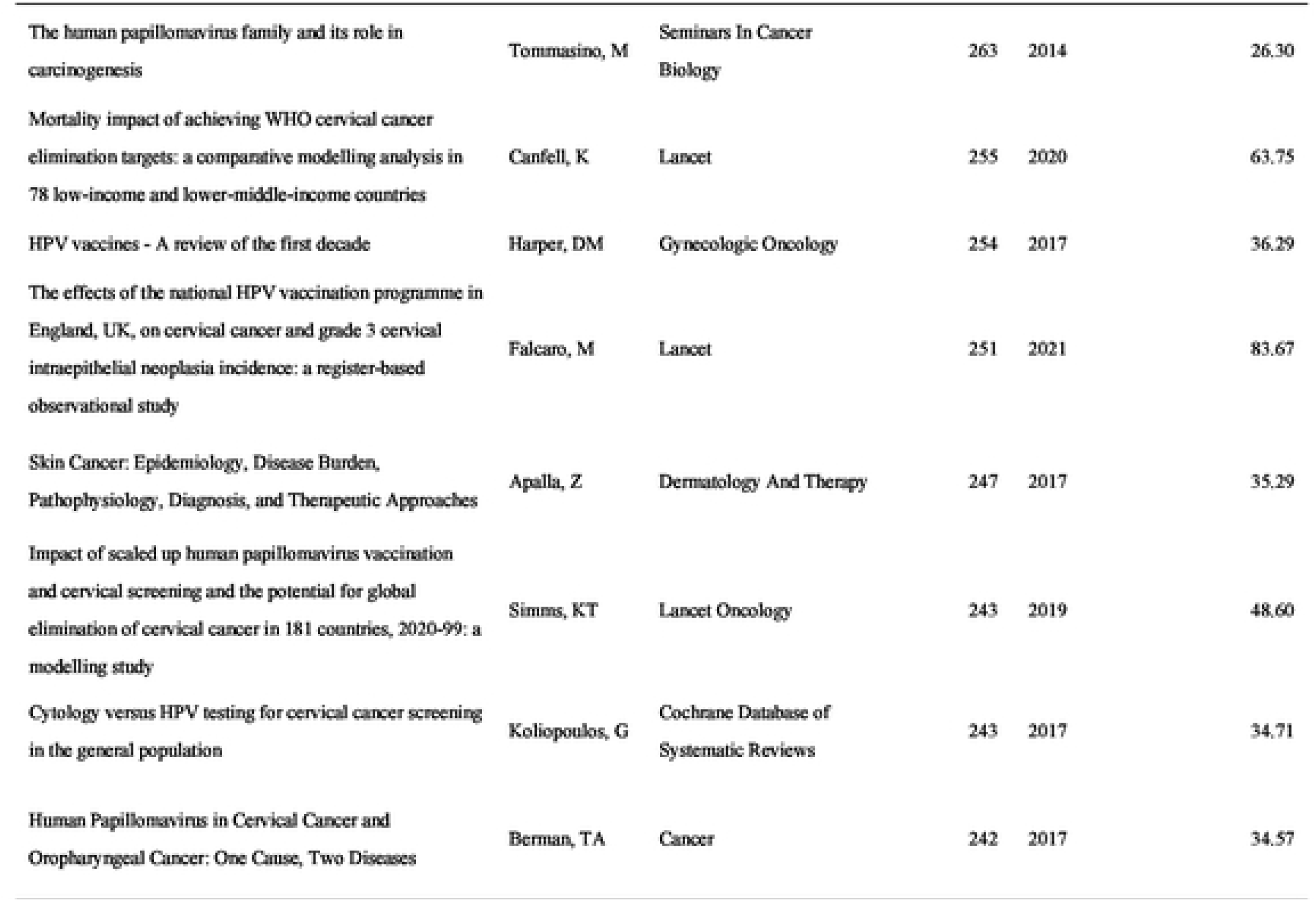
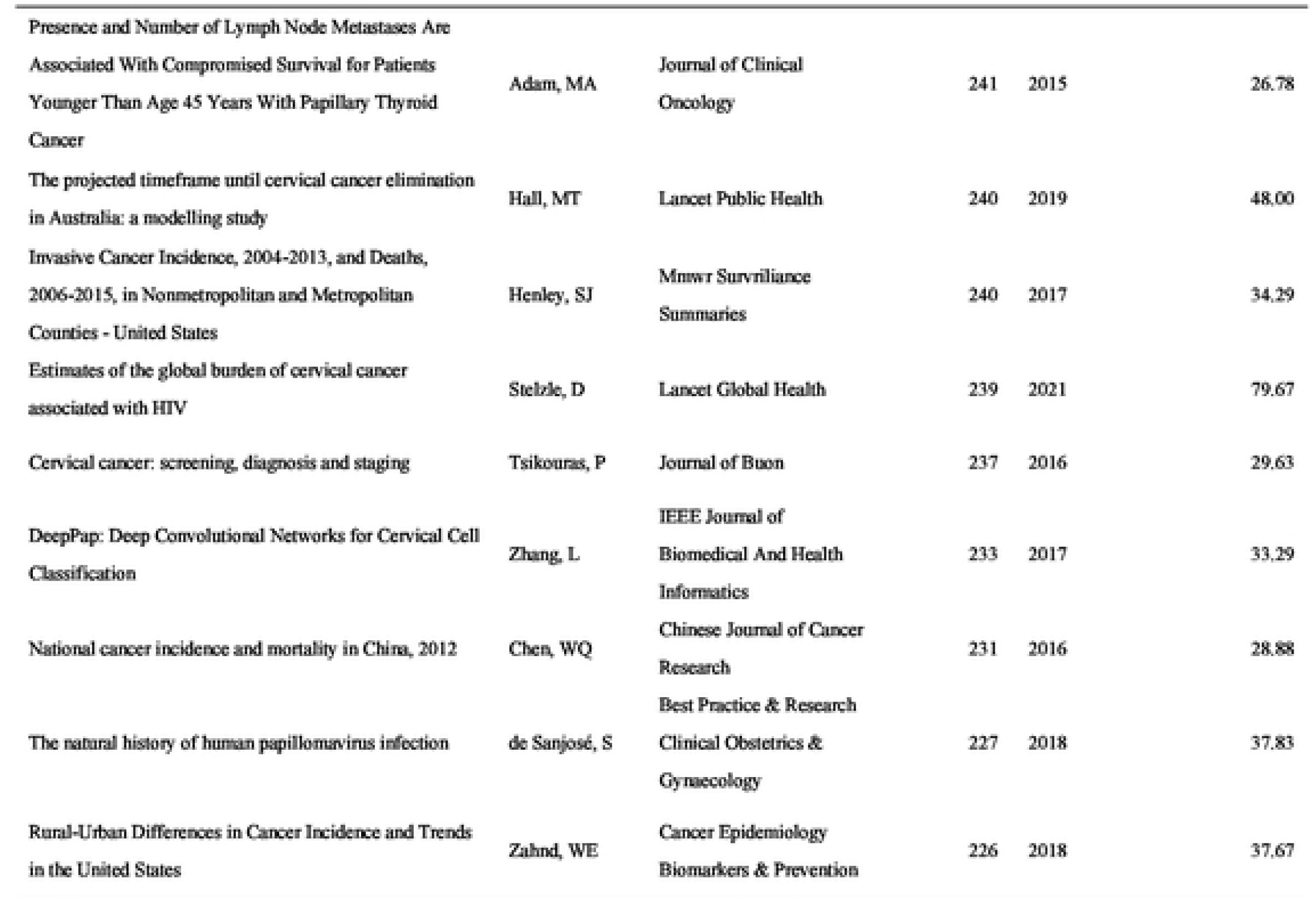
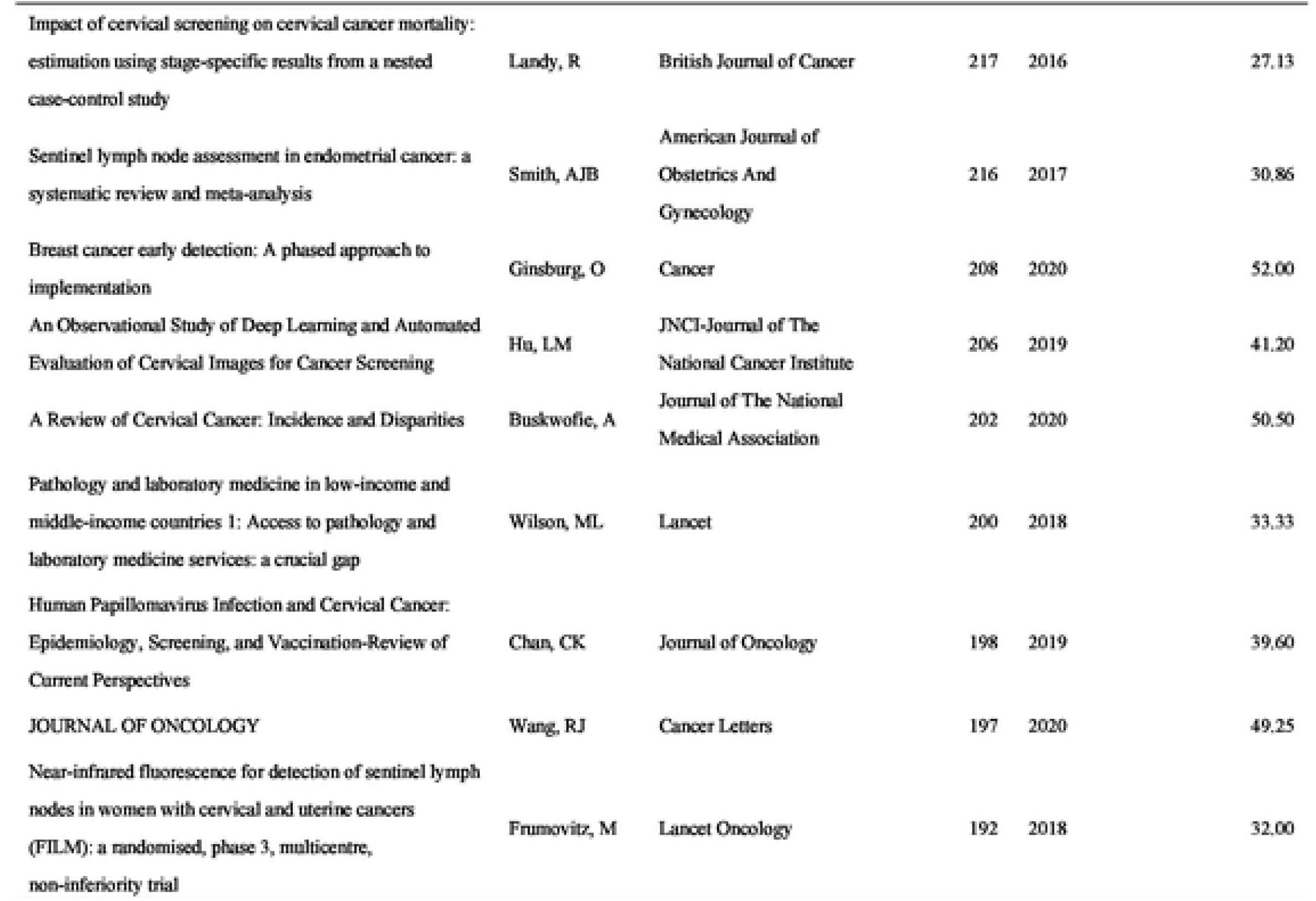
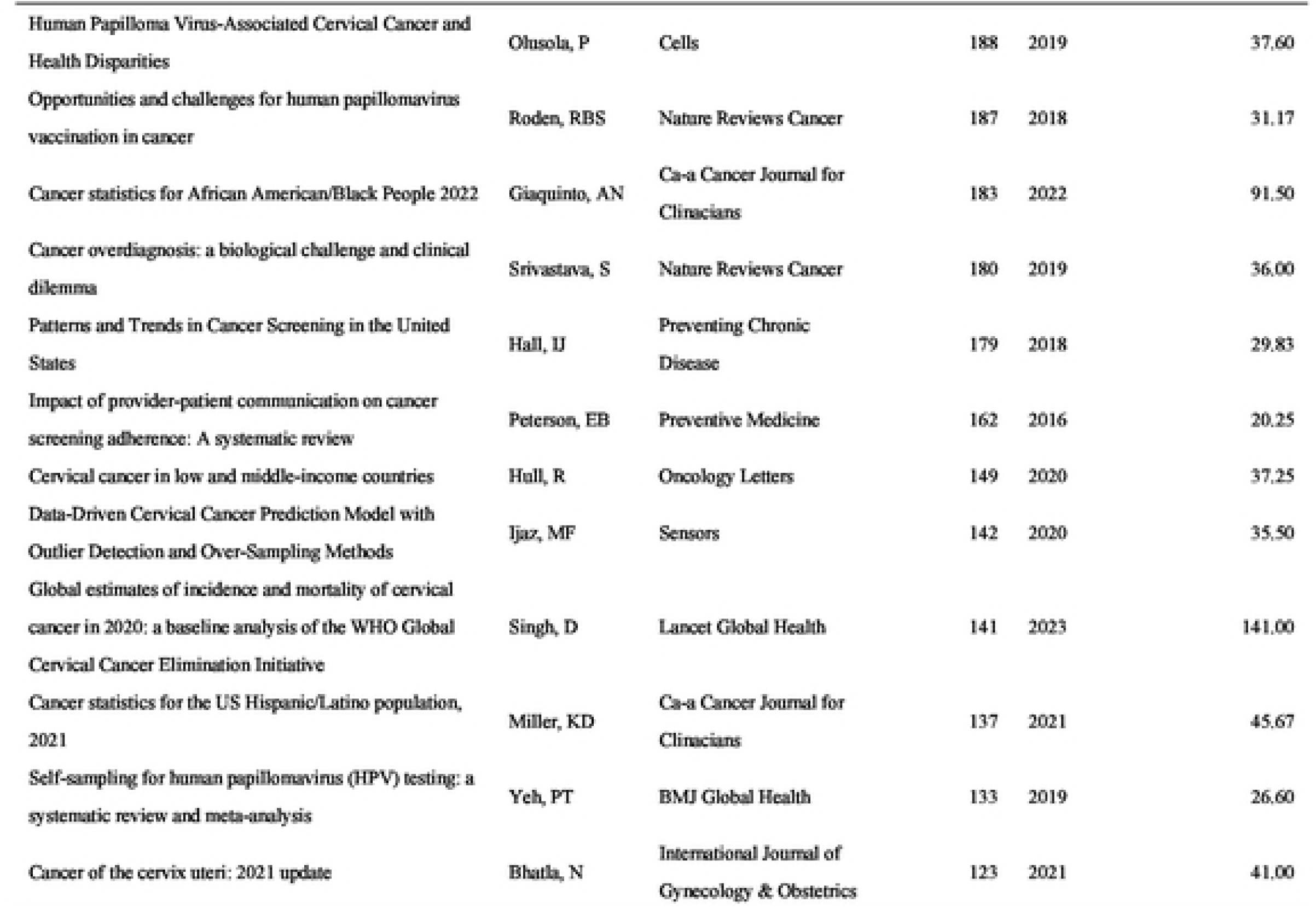
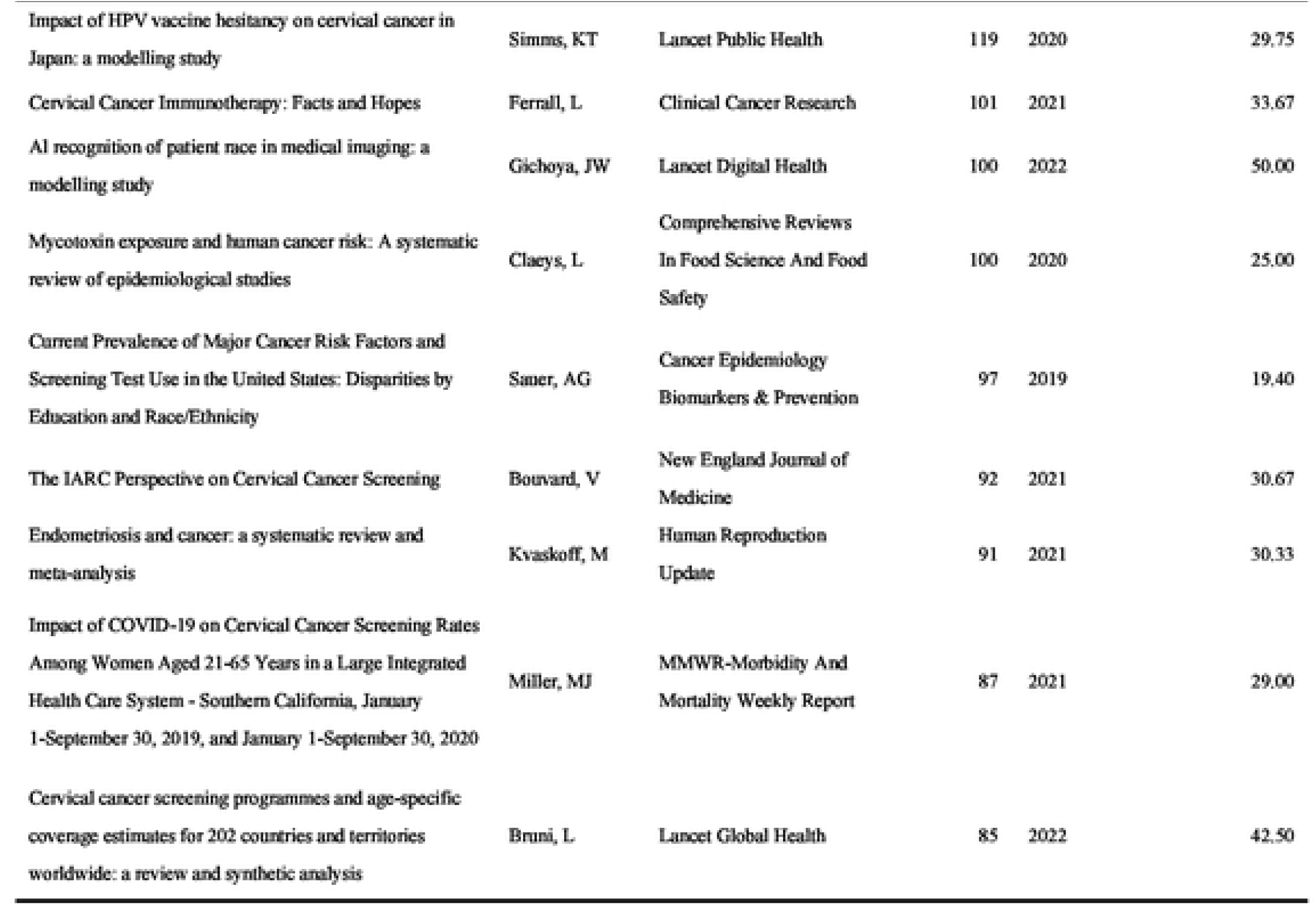
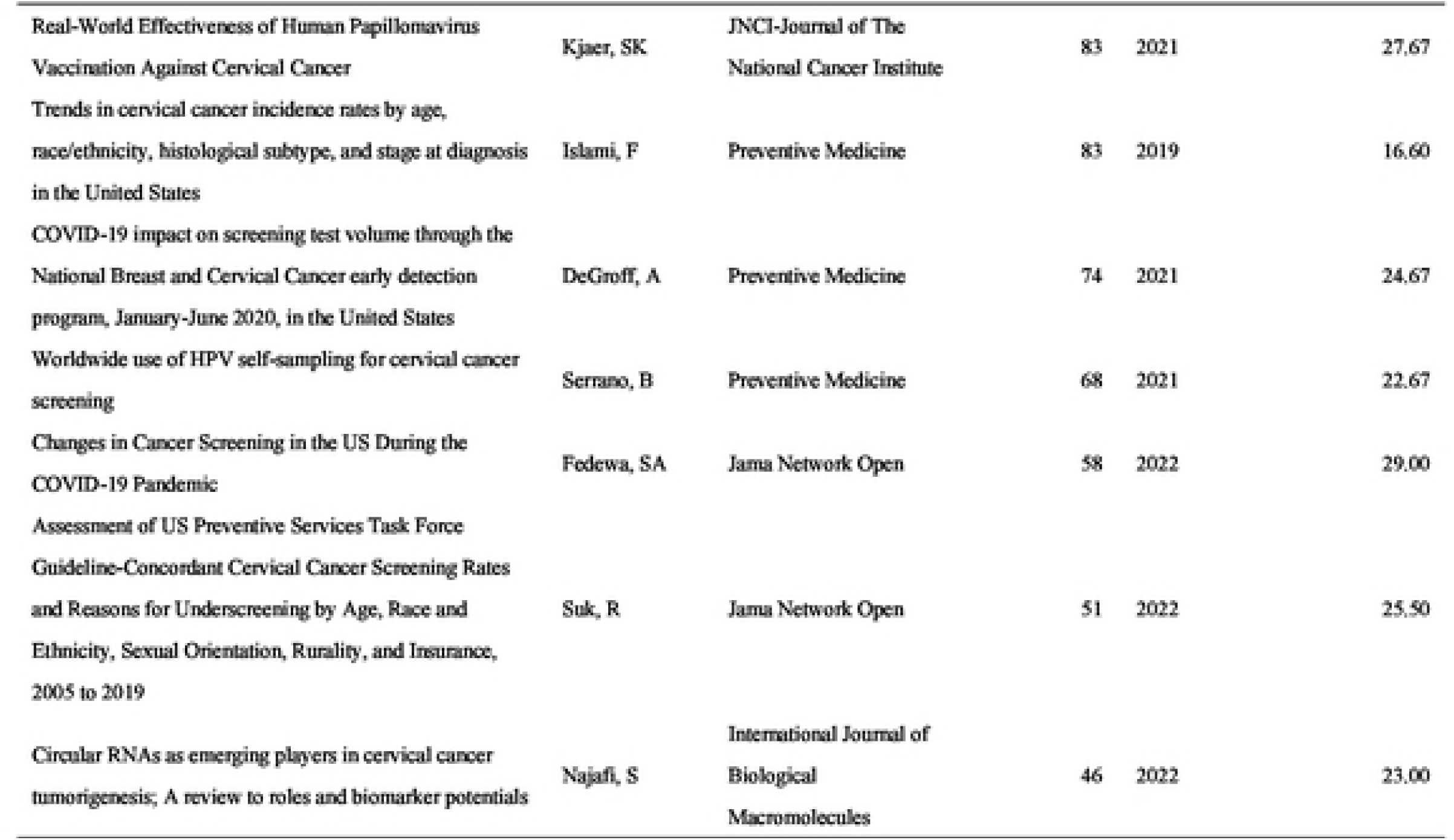
The top 100 cited papers in CCS.

### 3.2 Year of publication and citation

We depict their annual distribution, annual citation frequency distribution, and overall citation frequency distribution in a line graph (Figure 1). The top 100 most cited papers were written between 2013 and 2023. Fourteen of the top 100 most-cited papers were published in 2017, 2018, and 2019. The paper with the highest total number of citations was published in 2015 (14,567 citations) and had the greatest impact on CC research.

**Fig. 1.**
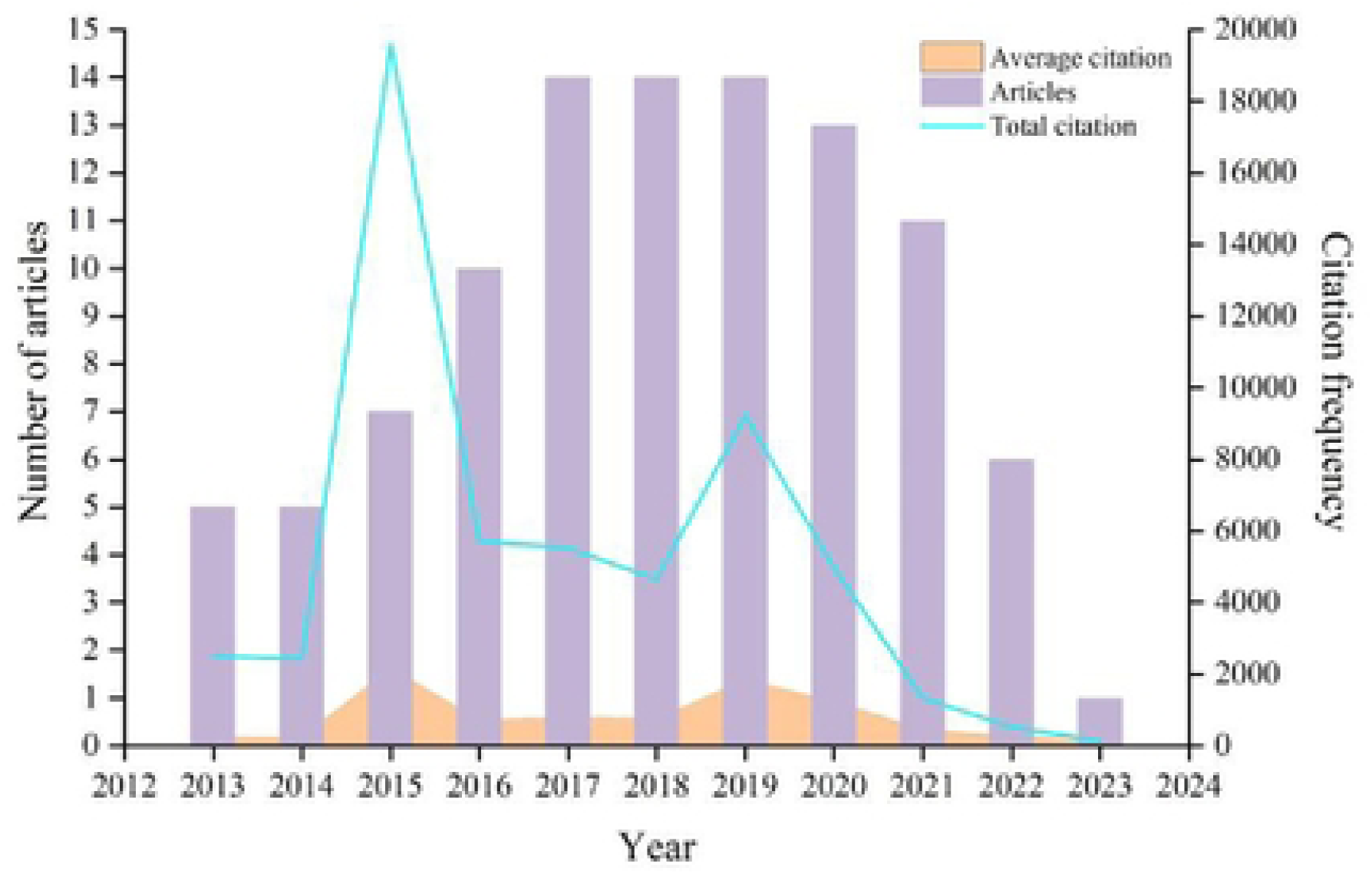
Year of publication and citation

### 3.3 Author and co-author analysis

There was active cooperation among some of the author clusters. (Fig. 2) The top 100 cited publications were contributed by 264 different authors. Jemal, Ahmedin authored the greatest number of papers (N = 12), followed by Sigel, Rebecca L (N = 8). Utilizing Jemal, Ahmedin as its core, the network’s total connection strength is 292, with the largest group of linked objects consisting of 184 items. In Figure 2, we can see that there is a direct linkage between Jemal, Ahmedin, and Sigel, Rebecca L. This means that they have a collaborative relationship, as a team to conduct their research.

**Fig. 2.**
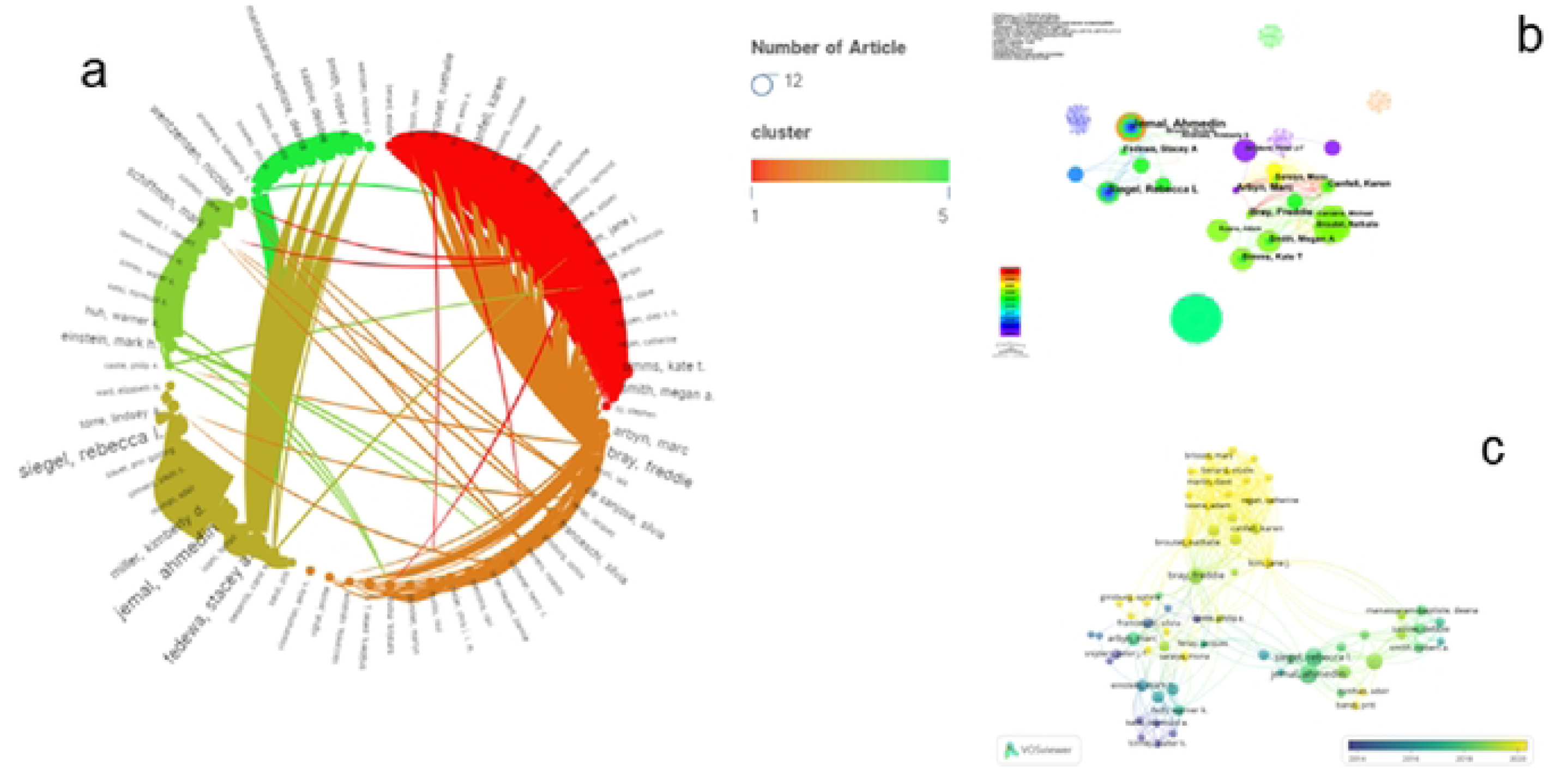
Network visualization of authors that contributed to the top 100 cited papers

### 3.4 Distribution of countries/regions and institutions

#### Countries/regions

A total of 240 countries/regions did research on the top 100 cited papers. The USA contributed the most publications (N = 70), followed by the FRANCE (N = 25) and the ENGLAND (N = 19). Seven main clusters were formed by all the countries/regions. The USA established the largest national cooperative network, encompassing 70 countries/regions, followed by France, which covered 25 countries/regions. The bibliometric map highlighted the close relationship between countries/regions (Fig.3). Collaborative efforts across countries/regions resulted in thicker line segments between nodes. In recent years, Japan and India have had the highest publication numbers. The bibliometric map highlighted the close relationship between countries/regions.

**Fig. 3.**
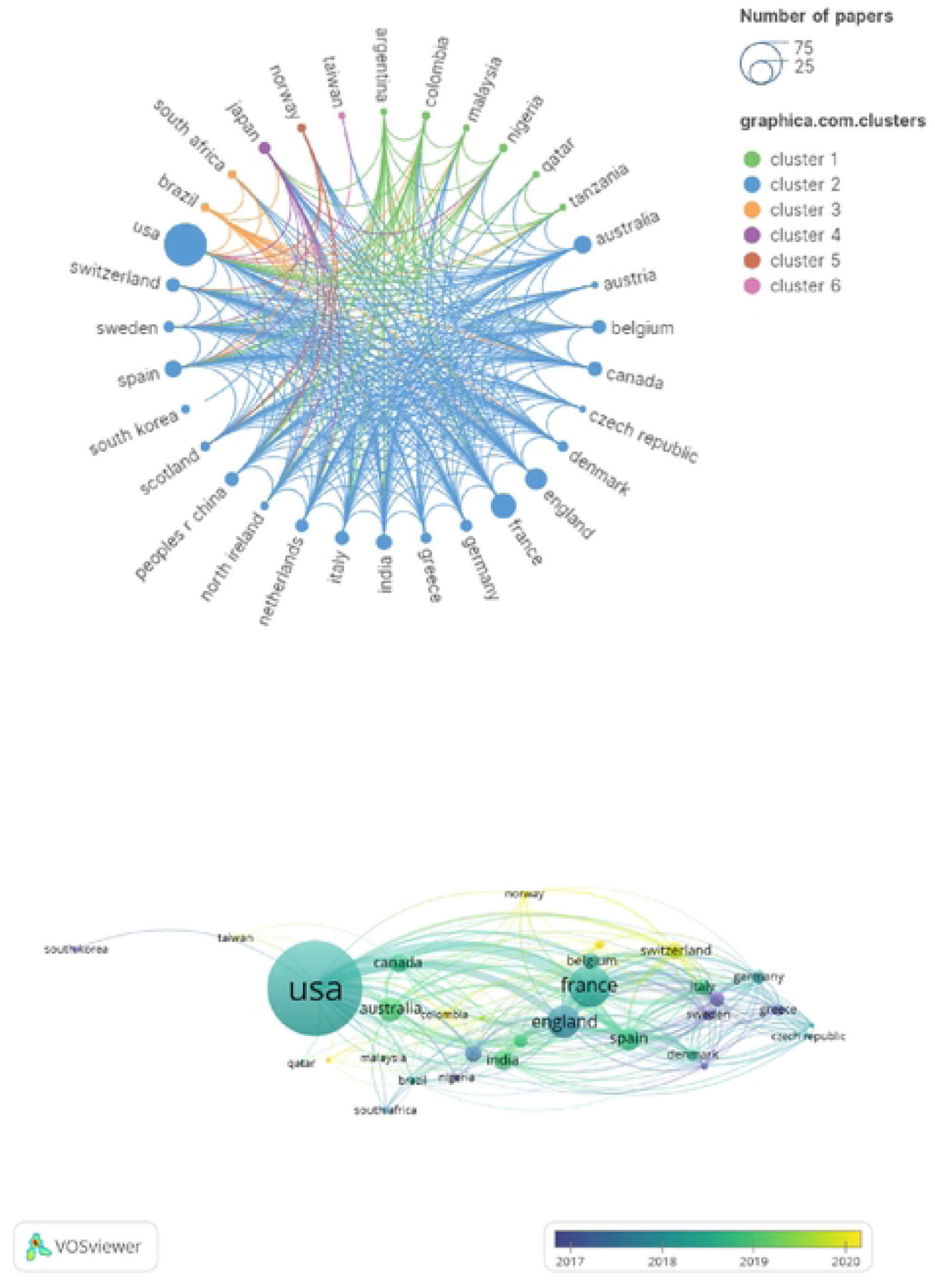
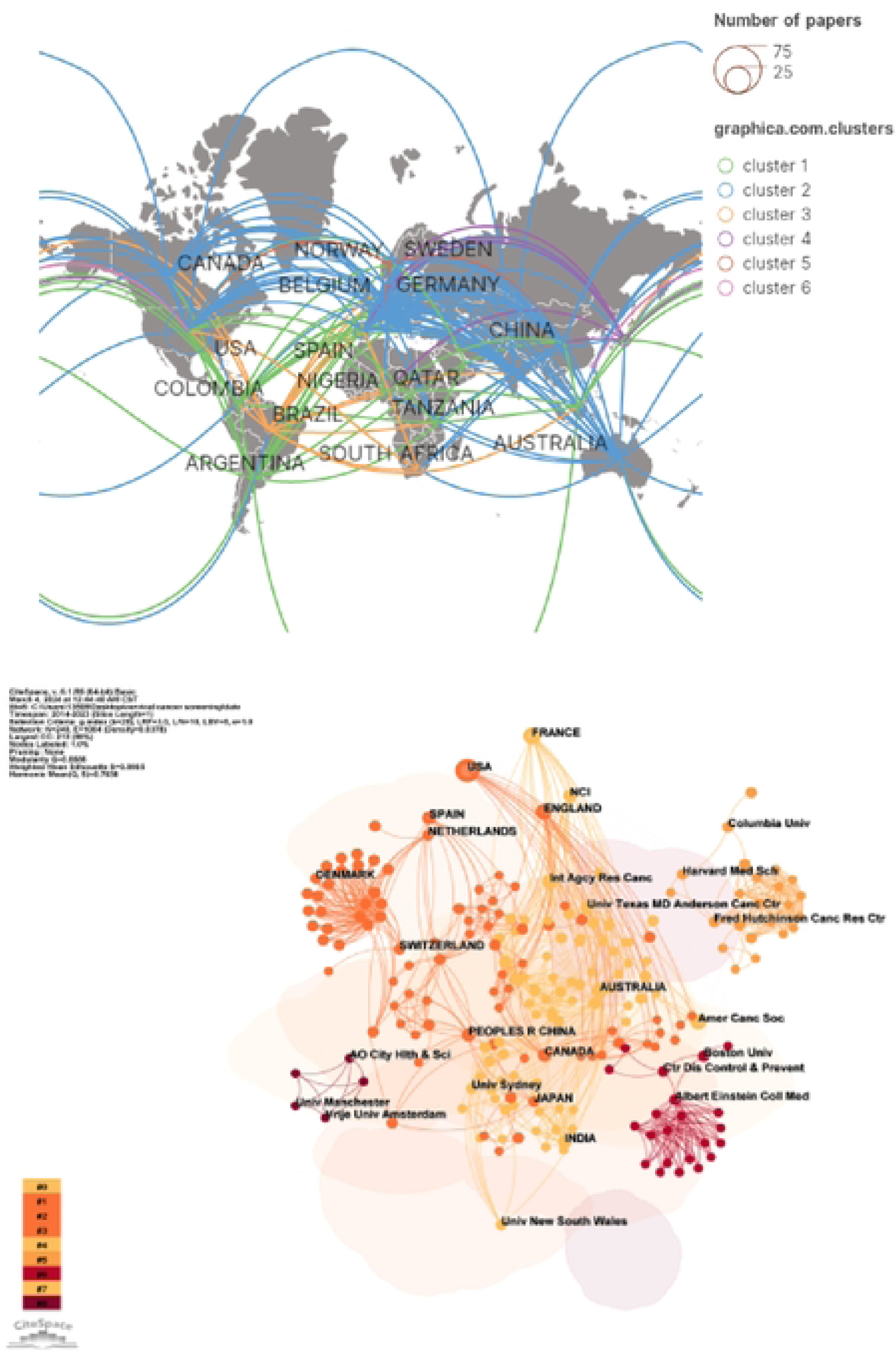
Networks showing the collaboration among countries in the top 100 cited papers

#### Institutions

Of the included studies, Int Agcy Res Canc & Amer Canc Soc contributed the most papers (N = 17), followed by NCI (N = 10), which were the institutions with more than 10 publications. Also, Univ Texas MD Anderson Canc Ctr (N = 7), Ctr Dis Control & Prevent, and Univ Sydney (N = 6) had the most publications, and Amer Canc Soc had the most citations (N = 24157), followed by Int Agcy Res. Canc (N = 7429) and NCI (N = 2046). Int Agcy Res Canc and Amer Canc Soc are institutions that have published more articles in recent years. (Fig.4)

**Fig. 4.**
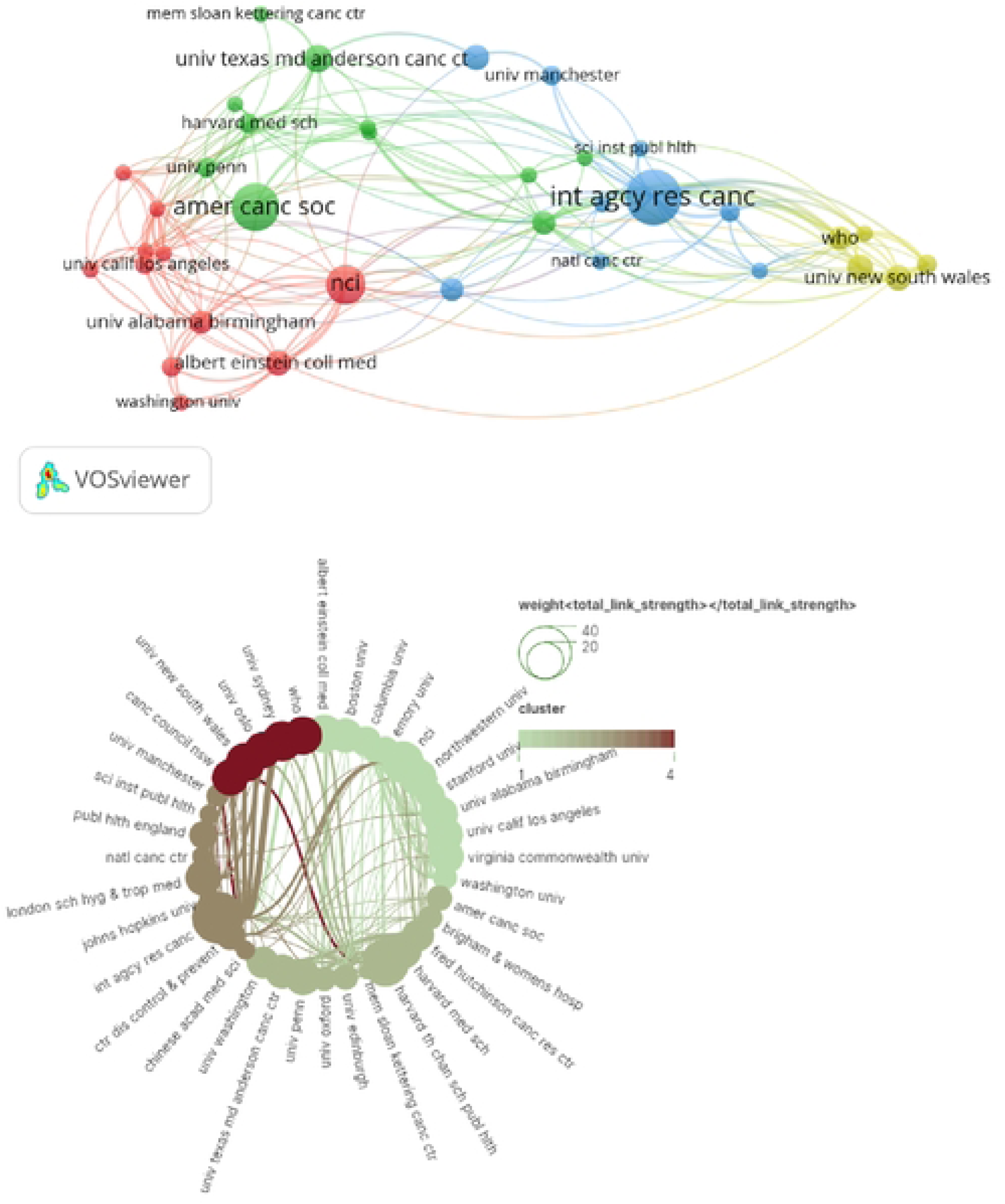
Network visualization of the institutions that contributed to the top 100 cited papers

### 3.5 Journal

The dual-journal map overlay shows the three main citation pathways (Figure 5), with papers published in Medicine/ Medicine/ Clinical/ Sports/ Neuro/ Immunity journals citing Genes/ Biology/ Genetics and Surgery/ Rehabilitation/ Sports. paper. This conclusion can be used as a reference for new scholars engaged in CCS research.

**Fig. 5.**
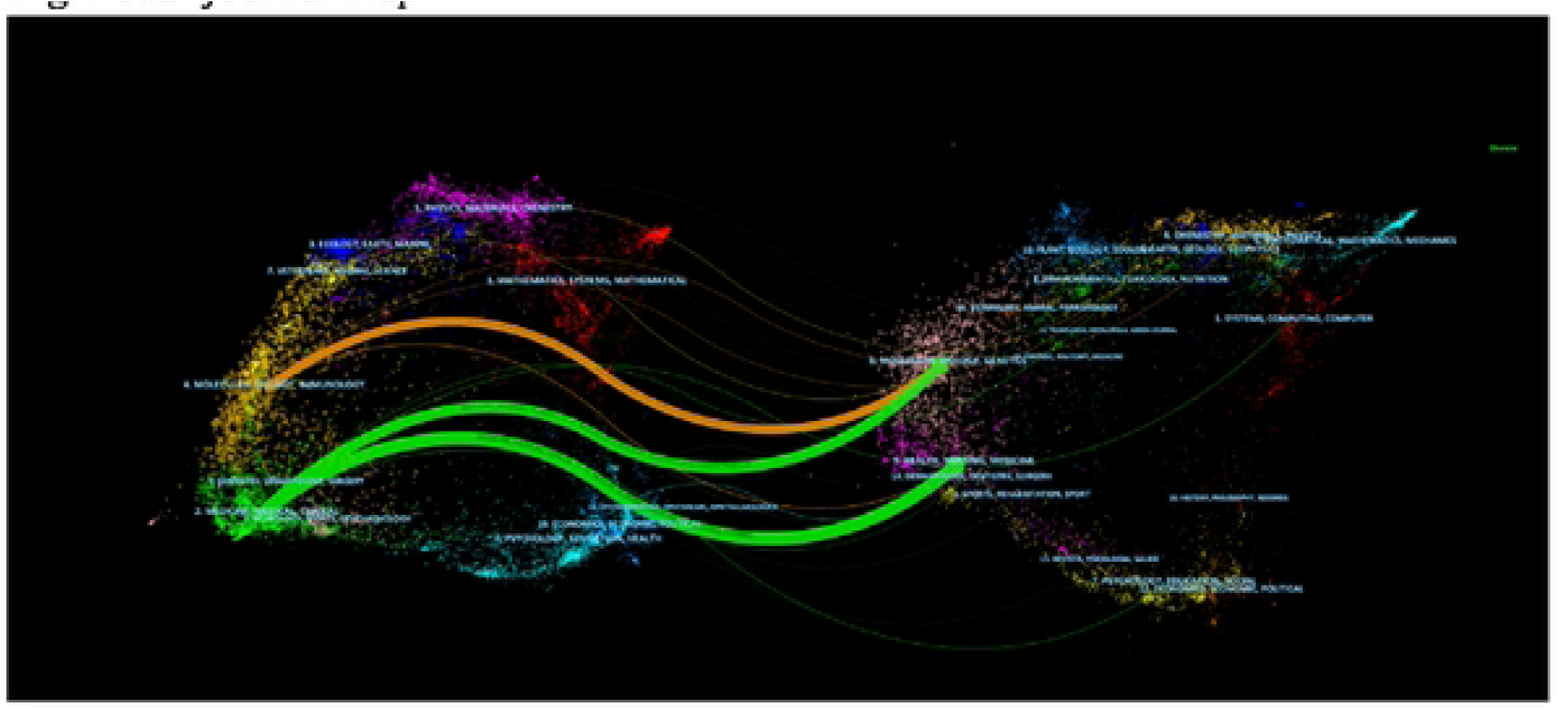
dual-journal map

### 3.6 Keywords co-occurrence, clusters, and bursts

A total of 242 keywords were identified in the top 100 cited papers, with a total of 997 connecting lines between keywords. We excluded keywords such as place names and then evaluated the data shown in the graphs. The most common keywords were cervical cancer (N = 34), women (N = 21), human papillomavirus (N = 16), colorectal cancer (N = 16), and breast cancer (N = 15). Among them, human papillomavirus (N = 16) was an important causative factor for cervical cancer. Colorectal cancer (N = 16), breast cancer (N = 15), and lung cancer (N = 9) were strongly associated with cervical cancer. Human papillomavirus vaccination (N = 10) is an effective means of preventing cervical cancer (Table 3). In other areas, The United States (N = 16) and Africa (N = 2) were the two most frequent countries. Pelvic radiation therapy (N = 4) as a treatment for advanced cervical cancer.

**Table 3.** Top 10 co-occurring keywords in the top 100 cited papers on CCS.

| Rank | Keyword | Occurrences |
| --- | --- | --- |
| 1 | cervical cancer | 34 |
| 2 | women | 21 |
| 3 | human papillonmavirus | 16 |
| 4 | United states | 16 |
| 5 | colorectal cancer | 16 |
| 6 | Breast cancer | 15 |
| 7 | Follow up | 10 |
| 8 | Intraepithelial neoplasia | 10 |
| 9 | human papillonmavirus vaccination | 10 |

Clustering makes it easy to determine the distribution of research content on a given topic. The smaller the cluster ordinal number, the more nodes it contains, indicating that the more keywords it contains, the hotter the clustering research. These keywords are further subdivided into 13 clusters, of which the first 10 clusters are as follows (Fig. 6): cluster 1 (cancer statistics), cluster 2 (cervical screening), cluster 3 (hpv vaccine), cluster 4 (human papillomavirus testing), cluster 5 (systematic review), cluster 6 (three-layer perceptron), cluster 7 (cervical cancer tumorigenesis), cluster 8 (cancer screening), cluster 9 (ASCAP).

**Fig. 6.**
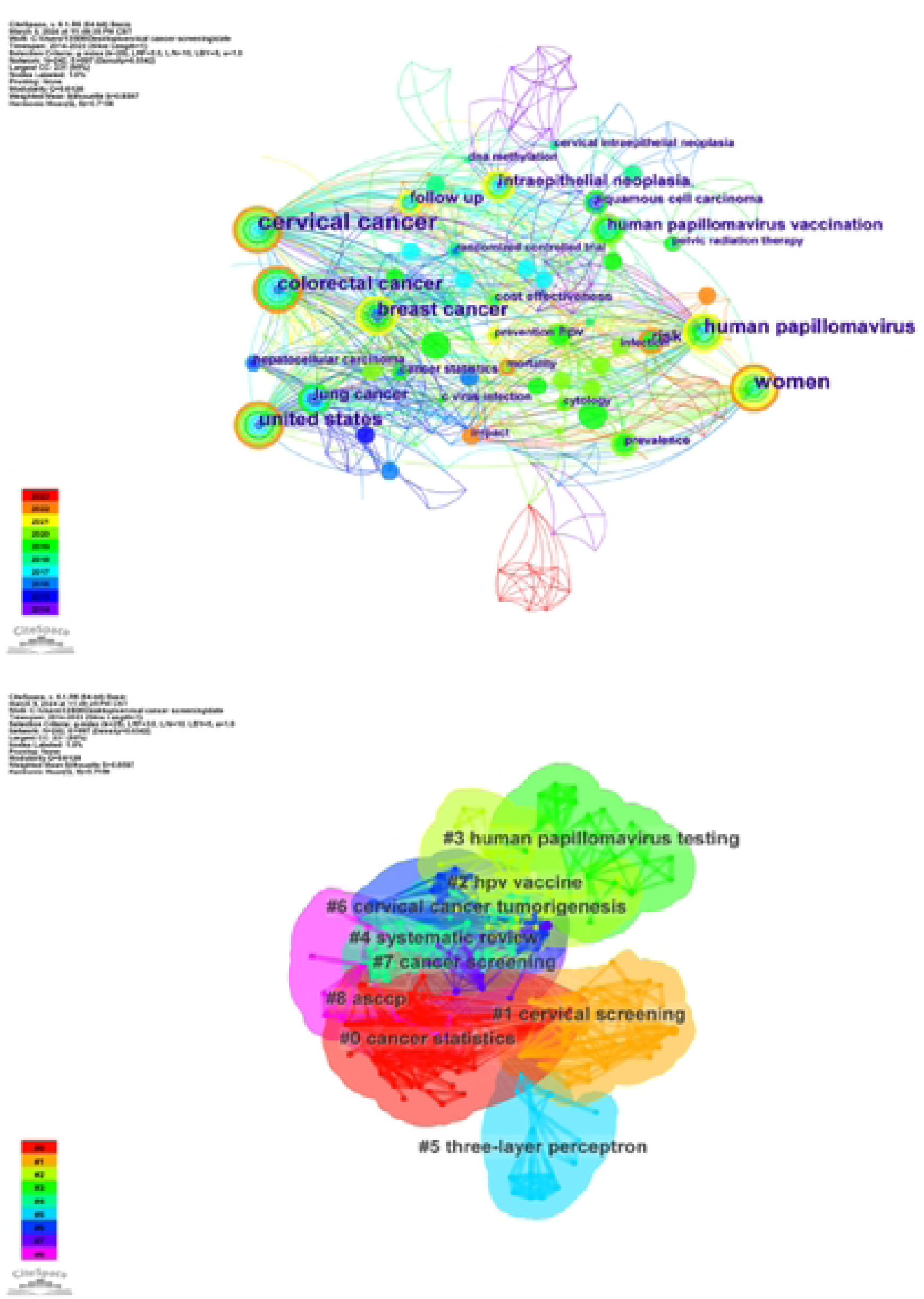
Network visualization keywords and clustering

We created a keyword timeline chart to analyze trends in keyword research in recent years (Fig. 7). Back before 2014, researchers realized that long-term HPV infection can lead to cervical cancer, and HPV, cervical cancer screening, and HPV vaccine to prevent cervical cancer have been hot research topics in recent years. In recent years, researchers have studied the development of cervical cancer and the safety of treatment from cytology and molecular. The results of cluster analysis suggest that the safety and efficacy of cervical cancer prevention, screening, and treatment may be the focus and trend of future research on CCS.

**Fig. 7.**
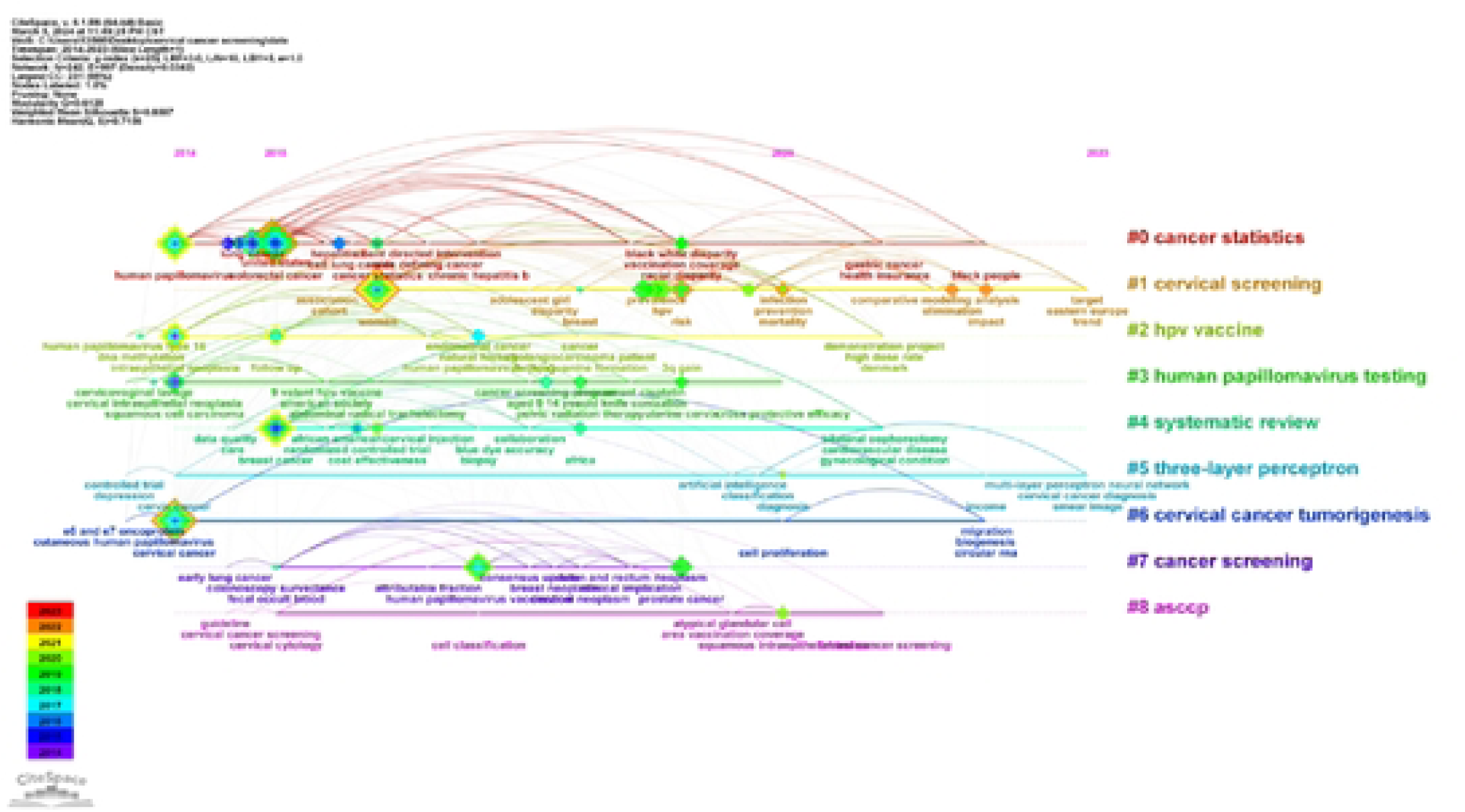
Keyword clustering analysis of the top 100 cited papers changes by year

### 3.7 Burst

We created keyword emergence charts to illustrate the hot trends in research for cervical cancer screening in recent years (Fig. 8). Initially, there was interest in the relationship between breast, colorectal, and lung cancer and cervical cancer. Human papillomavirus was the longest studied of all the keywords. In recent years, researchers have been focusing on the diagnosis, infection mortality, and risk.

**Fig. 8.**
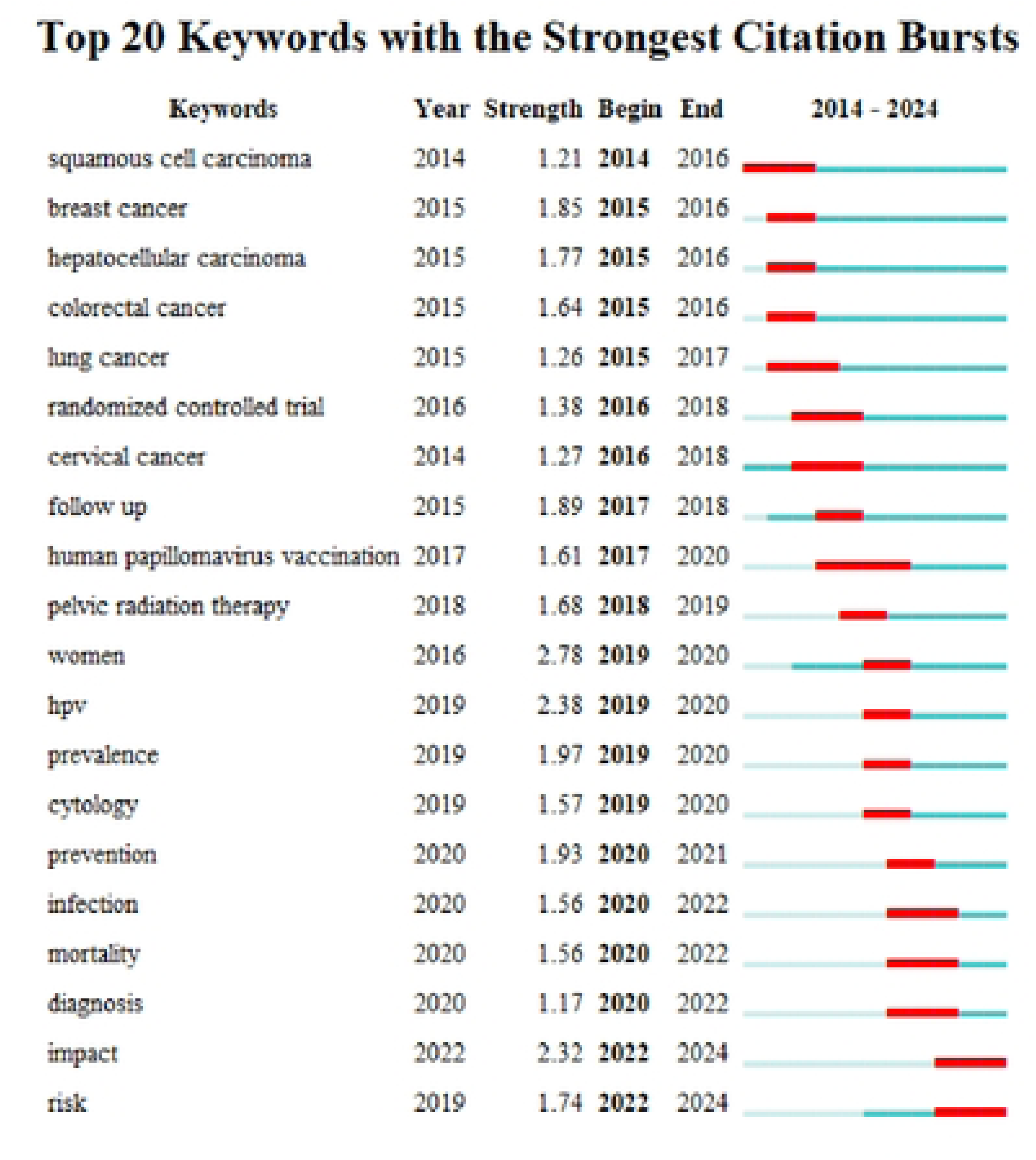
Network visualization of the keywords with the strongest citation bursts of the top 100 cited papers

Infection and diagnosis have become a new research topic, indicating that researchers are more and more focused on the prevention and diagnosis of cervical cancer. The results of cluster analysis suggest that prevention of infection and early diagnosis of cervical cancer may be the focus and trend of future research on CCS.

### 3.8 Co-cited articles and co-cited reference cluster

Typically we use co-citation relationships to determine the degree of connectivity between different publications, which is said to occur if two or more articles are cited in one or more articles at the same time. Therefore, we create cluster network diagrams and co-citation relationships via citespace (Fig. 9). According to the clustering diagram (Q = 0.8602, S = 0.9525), the quality of the clustering diagram is generally considered to be better when Q > 0.3 and S > 0.5. Therefore, the results of the clustering diagram in this study are convincing. Figure 11 depicts the evolution of cluster analyses over several years of co-cited papers and co-cited references for the top 100 cited documents. We used CiteSpace software to calculate the citations with the highest burst intensity over the last 10 years. Figure 10 lists the cited papers[19–42].

**Fig. 9.**
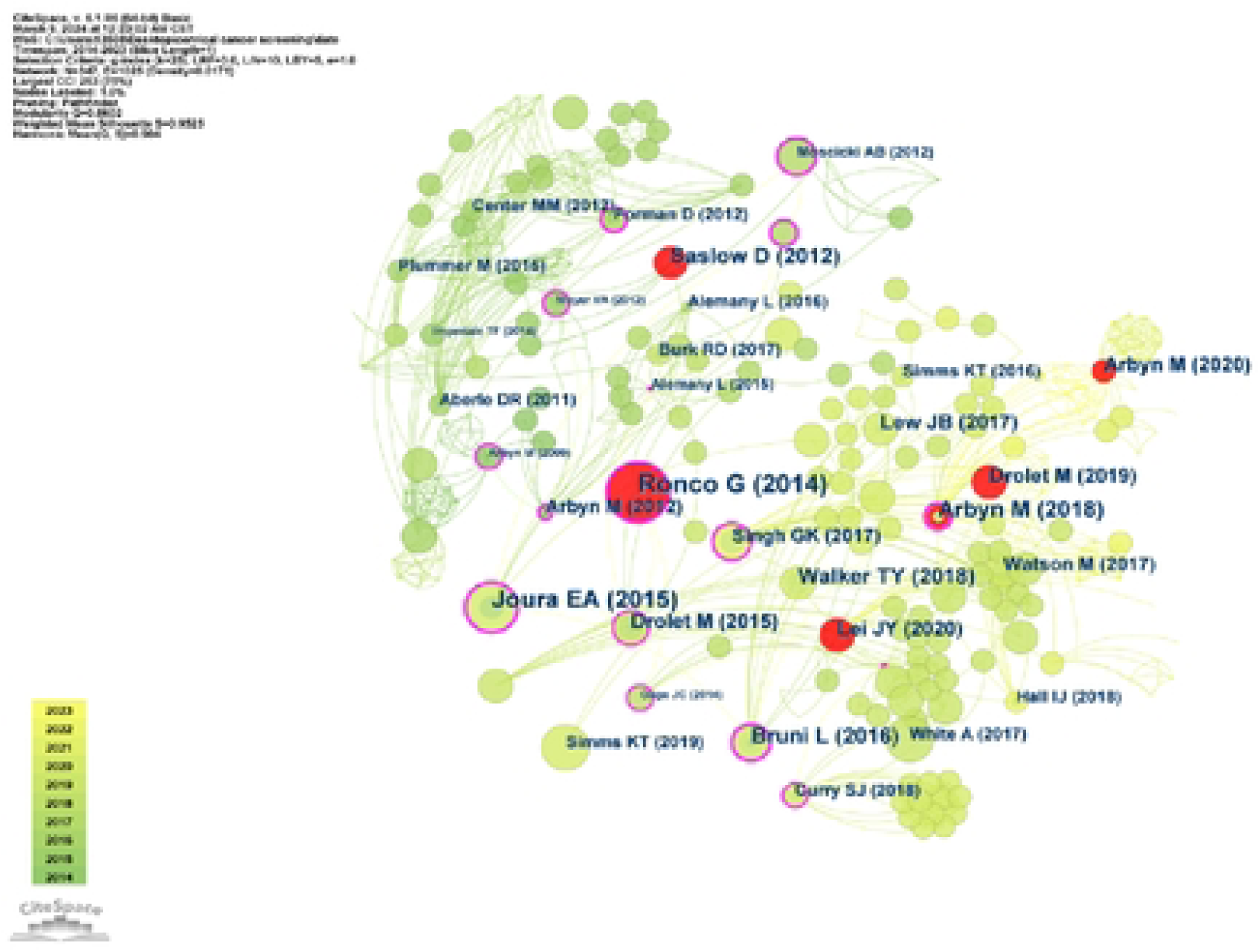
Co-cited articles and co-cited reference analysis of the top 100 cited papers

**Fig. 10.**
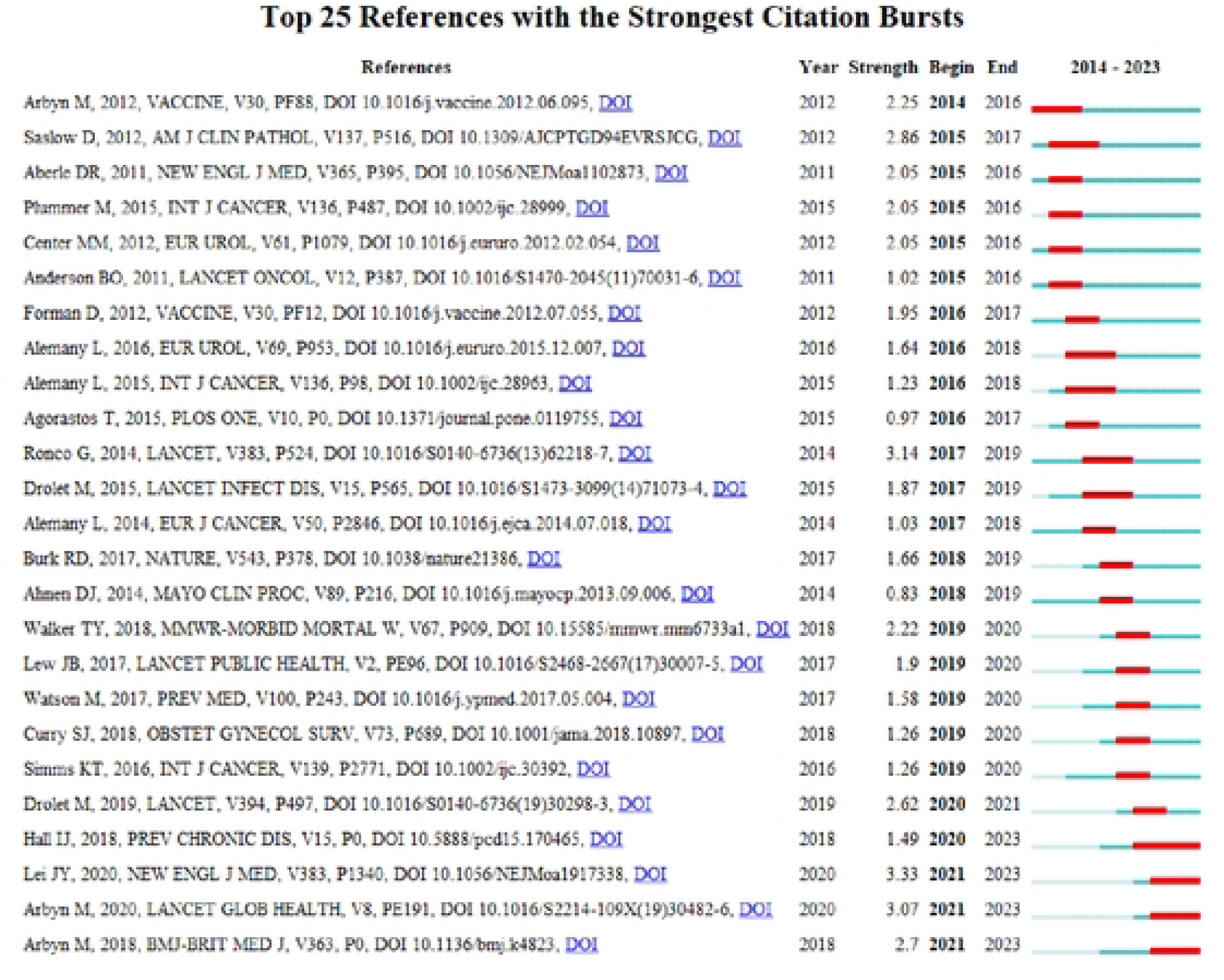
Top 25 references with the strongest citation bursts of the top 100 cited papers

**Fig. 11.**
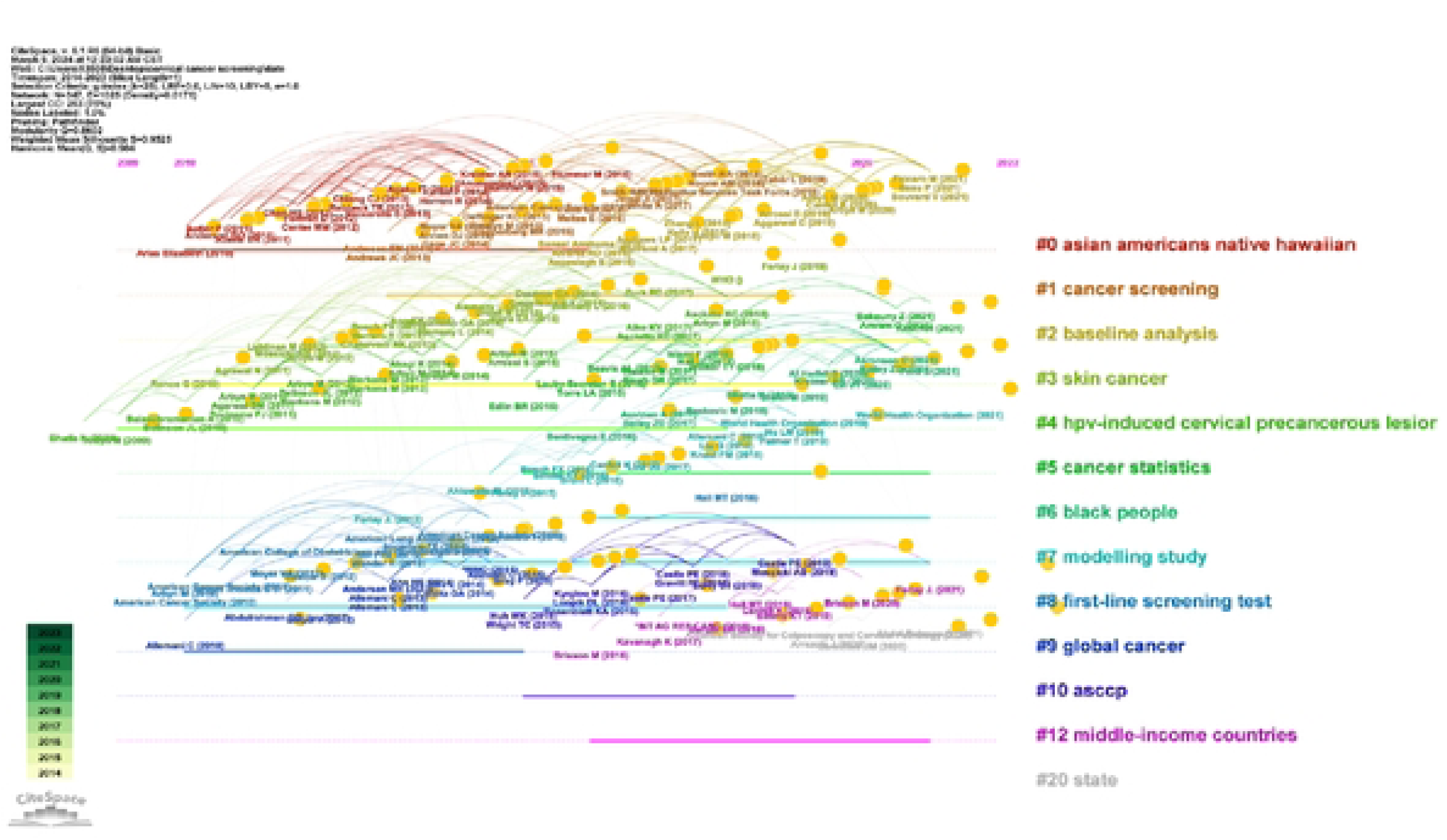
Co-cited articles and co-cited reference cluster analysis of the top 100 cited papers changes by year

Of all the discourses on CCS, a multicenter study by Ronco, G. et al. showed the importance of HPV-based or liquid-based cytology for early CC detection. Arbyn, M tested hr-HPV women for cervical intraepithelial neoplasia grade 2 or worse by performing hrHPV testing as well as colposcopy and biopsy and therefore concluded that hrHPV testing is important in the early diagnosis of CC. Hall, MT used a modeling study to estimate the age-standardized incidence of CC in HPV-vaccinated women a few years later and is on track to reach a target of 4 new cases per 100,000 women per year in the future. In conclusion, most trials have shown the importance of early screening for the diagnosis of CC and the availability of a CC vaccine to help prevent CC.

**Analysis**

## Discussion

Over the past nearly two decades, researchers have made many efforts and achieved promising results in the early diagnosis and prevention of CC, but barriers remain and future ones need to be broken through.

### General information from the top 100 cited papers

In this study, we summarised the 100 most cited papers screened by CCS. Using rigorously standardized bibliometric and visual analysis methods, we derive global trends regarding the most frequently cited literature in the field of CCS research. Based on specific analyses and reliable metrics, bibliometric analyses can be analysed to summarise the characteristics of published articles, and assessing the most cited papers can provide medical researchers in the field with both important information about the focus of this research direction and future research directions.

In terms of the types of articles selected, most of the top 100 cited papers are original articles, mostly distributed between 2013 and 2023. The top 20 most cited papers account for approximately 80% of all citations in the top 100 most cited papers. Ronco, G was the main contributor to the most influential article, indicating that their research is a potential hotspot in the field [17]. Siegel, RL et al. 2019 published “Cancer Statistics,2019” in Ca-a Cancer Journal for Clinicians and received the most citations [11]. By summarizing the morbidity and mortality of CC and the fact that CC has been effectively controlled in recent years, this article demonstrates that early diagnosis and prevention of CC have a huge impact on the diagnosis and treatment of the disease [43–46]. Recently Singh, D et al. published “Global Estimates of incidence and mortality of Cervical Cancer in 2020: A Baseline Analysis of the WHO Global Cervical Cancer Elimination Initiative” in Lancet Global Health addresses the dramatic changes in the global cervical cancer landscape today by assessing the extent of inequalities in global cervical cancer incidence and mortality to accelerate progress towards the WHO’s goal of eliminating CC [47].

The United States has the highest number of publications (N = 70), and in recent years, Japan and India have had the highest number of publications. This indicates that the United States has invested a lot of research in early screening and prevention of CC and has obtained significant results [44,46], and more countries are paying more attention to the prevention of CC [48–52].

### Limitations

This study also has several limitations as follows. Although Web of Science is the most commonly used database for literature searches, it may have some of its earlier publications missing. In order to perform an accurate search of the literature, we used a title keyword search method, which may result in them being accurate enough but may not be comprehensive enough.

## Conclusion

To our knowledge, this is the first bibliometric assessment of the most frequently cited literature on the early diagnosis of CC. The findings suggest that most of the important studies on CC were published in the last decade. Increasing attention is being paid to the early diagnosis and prevention of CC both nationally and internationally. Future research directions include universal access to vaccines, simplicity of early screening for CC, screening for risk genes relevant to clinical studies of CC, and clinical trials of treatments.

## Data Availability

All files are available from the database.

## Author contributions

LL conducted this study, produced the first draft, and refined it. Literature search, retrieval and data collection were conducted by CSC, SCX and CC. Data visualisation and graphical interpretation were carried out by CC and CSC.SL and YB were instrumental in providing significant support or funding. All authors participated and approved the final paper before submission.

## Declarations

The authors declare that the study was conducted in the absence of any commercial or financial relationships that could be interpreted as potential conflicts of interest.

Informed consent is not applicable.

## Informed consent

Not applicable

## Ethical review

This article is a visual analysis of a published article to which ethical review does not apply.

## Abbreviations

CCS: cervical cancer screening
CC: cervical cancer
IF: impact factor
RCT: randomized controlled trial
WoSCC: Web of Science Core Collection.

